# Evolution of SARS-CoV-2 seroprevalence and clusters in school children from June 2020 to April 2021 reflect community transmission: prospective cohort study *Ciao Corona*

**DOI:** 10.1101/2021.07.19.21260644

**Authors:** Agne Ulyte, Thomas Radtke, Irene A. Abela, Sarah R. Haile, Priska Ammann, Christoph Berger, Alexandra Trkola, Jan Fehr, Milo A. Puhan, Susi Kriemler

**Affiliations:** Epidemiology, Biostatistics and Prevention Institute (EBPI), University of Zurich, Hirschengraben 84, 8001 Zürich, Switzerland; Institute of Medical Virology, University of Zurich, Winterthurerstrasse 190, 8057 Zürich, Switzerland; Division of Infectious Diseases & Hospital Epidemiology, University Hospital Zurich, Rämistrasse 100, 8091 Zurich, Switzerland; University Children Hospital Zurich, Steinwiesstrasse 75, 8032 Zürich, Switzerland; Institute of Medical Virology, University of Zurich, Winterthurerstrasse 190, 8057 Zürich, Switzerland; Epidemiology, Biostatistics and Prevention Institute (EBPI), University of Zurich, Hirschengraben 84, 8001 Zürich; Division of Infectious Diseases & Hospital Epidemiology, University Hospital Zurich, Rämistrasse 100, 8091 Zurich, Switzerland

**Keywords:** SARS-CoV-2, COVID-19, children, adolescents, school, SARS-CoV-2 symptoms, non-participation, participation rate, cohort

## Abstract

**Objectives:** To longitudinally assess severe acute respiratory syndrome coronavirus 2 (SARS-CoV-2) seroprevalence and clustering of seropositive children within school classes in March-April 2021 compared to June-July and October-November 2020. To examine the evolution of symptoms and the extent of under-detection of SARS-CoV-2 in children.

**Design:** Prospective cohort study of randomly selected schools and classes.

**Setting:** Schools remained open for physical attendance in Switzerland from May 2020 to the end of 2020/2021 school year. Lower school level (age range 7-10 years) and middle school level (8-13 years) children in primary schools, and upper school level (12-17 years) children in secondary schools were invited for SARS-CoV-2 serological testing in the *Ciao Corona* study in the canton of Zurich, Switzerland. Three testing rounds were completed in June-July 2020 (T1; after the first wave of SARS-CoV-2 infections), October-November 2020 (T2; during the peak of the second wave), and March-April 2021 (T3; after the second wave and with SARS-CoV-2 variants of concern becoming dominant). Parents completed questionnaires on sociodemographic information and symptoms.

**Participants:** 2487 children (median age 12 years, age range 7-17 years) recruited from 275 classes in 55 schools participated in the testing in March-April 2021; total of 2974 children participated in at least one of the 3 testing rounds.

**Main outcome measures:** SARS-CoV-2 serology results; clustering of seropositive children within classes; reported symptoms.

**Results:** The proportion of children who were SARS-CoV-2 seropositive increased from 1.5% (95% credible interval (CrI) 0.6% to 2.6%) in June-July 2020, to 6.6% (95% CrI 4.0% to 8.9%) in October-November, and to 16.4% (95% CrI 12.1% to 19.5%) in March-April 2021. By March-April 2021, children in upper school level (12.4%; 95% CrI 7.3% to 16.7%) were less likely to be seropositive than those in middle (19.5%; 95% CrI 14.2% to 24.4%) or lower school levels (16.0%; 95% CrI 11.0% to 20.4%). Children in the upper school level had a 5.1% (95% CI -9.4% to -0.7%) lower than expected seroprevalence by March-April 2021 than those in middle school level, based on difference-in-differences analysis. The ratio of PCR-diagnosed to all seropositive children changed from 1 to 21.7 (by June-July 2020) to 1 to 3.5 (by March-April 2021). Symptoms were reported by 37% of newly seropositive and 16% seronegative children. Potential clusters of 3 or more newly seropositive children were detected in 24 of 119 (20%) classes with a high participation rate, from which a median of 17 clusters could be expected due to random distribution of seropositive children within the classes. Clustering was lowest in middle and upper school levels. Retention rate in the cohort was high (84% of T1 participants attended T3). Among participants, supporting society and research were reported more commonly for participation than personal reasons. Fear of blood sampling was the most frequently reported reason for non-participation, reported for 64% of children.

**Conclusions:** By March-April 2021, 16.4% of children and adolescents were seropositive in the canton of Zurich, Switzerland. The majority of clusters of SARS-CoV-2 seropositive children in school classes could be explained by community rather than intra-class transmission of infections. Seroprevalence and clustering was lowest in upper school levels during all timepoints.

**Trial registration:** ClinicalTrials.gov NCT04448717.

**What is already known on the topic:**

- Transmission of SARS-CoV-2 in school setting largely followed community transmission in 2020.
- With implemented preventive measures, secondary attack rates were low and clustering of SARS-CoV-2 infections within classes and schools (outbreaks) were observed rarely.

**What this study adds:**

- With high community incidence and new variants of SARS-CoV-2, seroprevalence increased in school children between October 2020 – March 2021 in the canton of Zurich in Switzerland, and was higher in lower school levels.
- Most of the potential clusters of children who tested seropositive within classes could be explained by community rather than intra-class transmission of SARS-CoV-2, especially in middle and upper school levels.
- More children who tested seropositive in March-April 2021 were diagnosed and reported symptoms potentially related to SARS-CoV-2 infection more frequently than those who tested seropositive in June-July or October-November 2020.
- The most frequent reason for non-participation was fear of blood sampling (62% of children).

## Introduction

Many countries in the Northern hemisphere experienced a second or third wave of severe acute respiratory syndrome coronavirus 2 (SARS-CoV-2) infection pandemic in the winter of 2020/2021 [1]. High community incidence, as well as increasing prevalence of SARS-CoV-2 variants of concern (VOC), such as α (B.1.1.7), γ (P.1) and δ (B.1.617.2), made governments reconsider the stringency of public preventive measures. Various studies, including contact-tracing, seroprevalence and real-time polymerase chain reaction (RT-PCR) screening studies, have shown that the spread of SARS-CoV-2 infection within schools was not larger than in the surrounding community in 2020, and secondary attack rates were low, with only rare outbreaks [2–5]. However, less evidence exists if more contagious variants of SARS-CoV-2 changed the transmission in schools significantly.

Attendance of schools has been disrupted in many countries worldwide in 2020/2021, with half of the countries worldwide and in Europe closing schooling for at least 30 weeks from March 2020 to June 2021 [6], and schools fully or partially closed for 56 weeks in the USA, and for 27 weeks in the UK during this time. In contrast, schools were fully or partially closed only for 7 weeks in the same period in Switzerland, and fully open for physical attendance through the whole 2020/2021 school year. Nevertheless, by November 2020, only minimal clustering of seropositive children was observed within randomly selected schools and classes in the canton of Zurich, Switzerland, after 2-3 months of school attendance including 2-6 weeks of high community incidence [7]. These findings implied that intra-class transmission was not a major contributor to SARS-CoV-2 infections among the children. Only scarce information is available on the spread of SARS-CoV-2 infection in schools since December 2020, when α (B.1.1.7) variant of SARS-CoV-2 started spreading in Europe and other countries [8]. In Switzerland, at the beginning of March 2021 approximately 80% of SARS-CoV-2 infections were due to the α variant (see Appendix 1) [9].

A class is the basic unit within a school, in which children spend the majority of their school time. However, the spread of infections in classes has been only indirectly examined in contact-tracing studies or those analysing diagnosed SARS-CoV-2 infections in children [2,4,10,11]. Transmission within schools in 2021/2022 and subsequent school years remains relevant to study. Children below the age of 12 will be the last group to be offered vaccination. Preventive measures in school will likely need to be adjusted as, on the one hand, new variants spread, and on the other hand, more people, especially the vulnerable populations, become vaccinated.

The *Ciao Corona* study uniquely examines SARS-CoV-2 seroprevalence on the class, alongside school and district levels. The objectives of this study were to estimate the proportion of seropositive children and adolescents within school levels, cantonal districts and the region (canton of Zurich) by March-April 2021 and compare it to the previous measurements in June/July 2020 and October/November 2020; assess the association of seropositivity in March-April 2021 with reported symptoms and compare it to that from 2020; and assess the frequency and evolution of clustering of seropositive children within classes in schools over time. In addition, we examined the reasons for participation and non-participation in this cohort study, in order to address potential participation bias.

## Methods

The protocol with detailed study design [12] and results of the previous measurements of this longitudinal cohort study [7,13] are reported elsewhere. This study is part of the nationally coordinated research network *Corona Immunitas* in Switzerland [14]. The study follows a cohort of randomly selected schools and classes in the canton of Zurich, Switzerland. The canton has a population of 1.5 million linguistically and ethnically diverse inhabitants in both rural and urban settings, and comprises 18% of the Swiss population [15].

During the COVID-19 pandemic in Switzerland, physical attendance of schools was interrupted only for two months in March-May 2020. Children attended school continuously in 2020/2021 school year, except during the regular school holidays (6 weeks in July-August, 2 weeks in October, 2 weeks in December-January). Preventive measures, such as distancing in classrooms and teachers’ rooms, reduced mixing of classes, reduction of large group activities and school trips, were implemented with some variation between schools. All schools required ill children to stay home unless with very mild symptoms. Adults at school were required to wear masks from October 2020, secondary school children (upper school level, approximate ages 12-16 years) from November 2020, and primary school children in middle school level grades (9-13 years) from late January 2021.

School-specific contact tracing was implemented in school year 2020/2021, and contact-tracing was triggered by child or school personnel testing positive for SARS-CoV-2. Subsequent testing and quarantine recommendations depended on the specific situation. As a general rule, the whole class was quarantined when two or more infected children were detected in the class simultaneously. In case children were wearing masks, only close contacts were quarantined [16]. School-wide screening with RT-PCR testing was implemented in case of a suspected outbreak starting from February 2021, especially when detecting SARS-CoV-2 variant of concern (VOC). Repetitive pooled RT-PCR screening of children and personnel has been implemented voluntarily in a few schools in the canton and city of Zurich from April 2021, but did not affect schools enrolled in this study. Daily incidence of diagnosed SARS-CoV-2 cases between October 2020 and April 2021 in the canton of Zurich and Switzerland, and the proportion of VOC among them, is shown in Appendix 1.

### Population

As described in the previous publications [7,12,13], in May-June 2020, we randomly selected primary schools from all schools in the canton of Zurich, and matched the geographically closest secondary school (often, in the same school building). Number of schools invited in the 12 districts of the canton was proportional to population size. From 156 invited, 55 schools agreed to participate (Figure 1).

**Figure 1.**
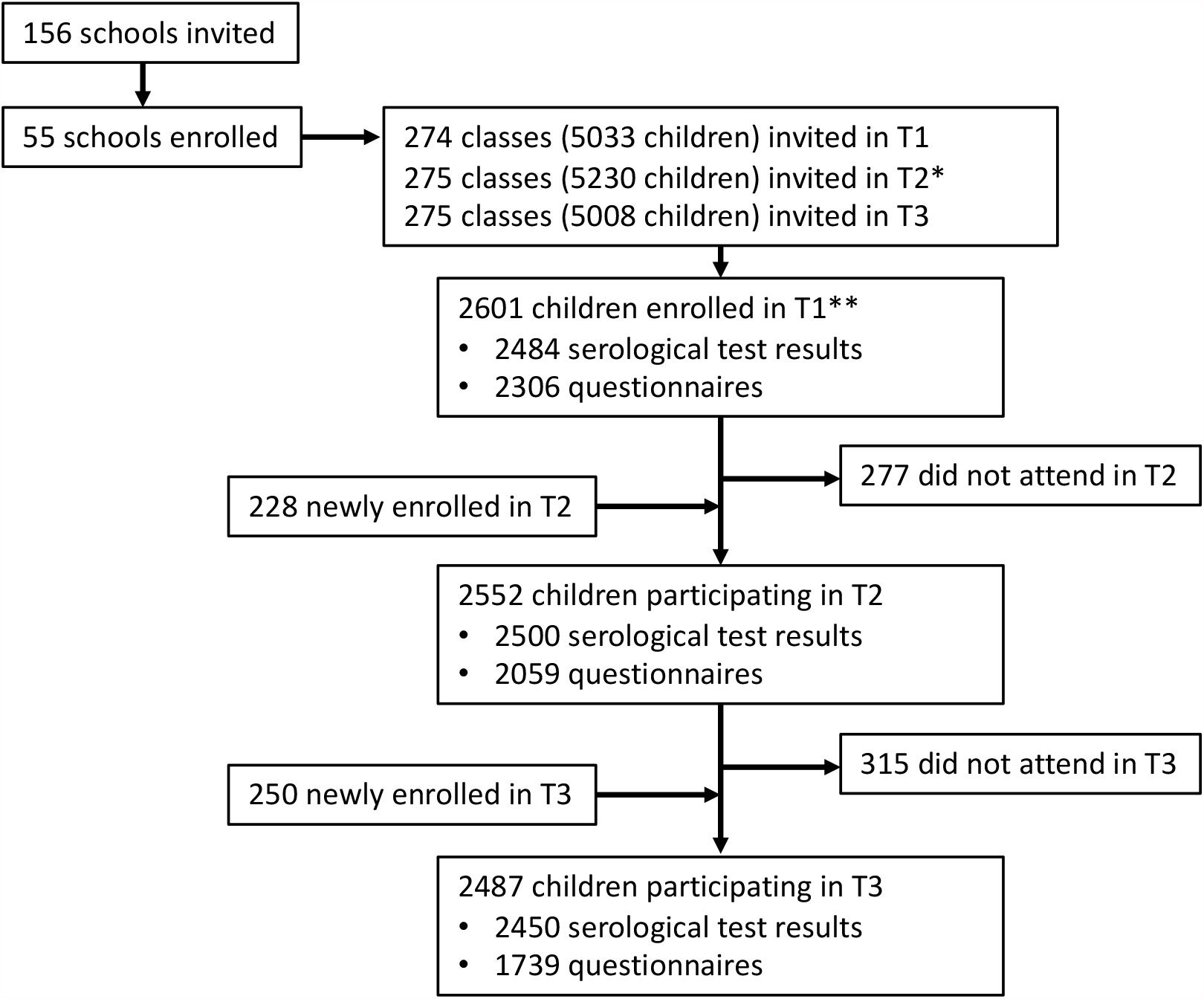
Flowchart of study participants. T1 – testing in June-July 2020, T2 – testing in October-November 2020, T3 – testing in March-April 2021. *Newly enrolled* were not tested in the previous round; *did not attend* means they did not attend subsequent round. ^*^ Some classes were split or rearranged after the summer break. ^**^ 16 of these children were enrolled from late August to early September 2020 (10 serological results, 16 questionnaires available).

We randomly selected classes within participating schools, stratified by school level: grades 1 to 2 in lower level (attended by 6 to 9-year-old-children), grades 4 to 5 in middle level (attended by 9 to 13-year-old children) and grades 7 to 8 in upper school level (attended by 12 to 16-year-old-children). Invited grades were selected to ensure that the same cohort of children will remain in the classes until April 2021 (children in grades 3, 6 and 9 often change the class and school in the next year). For this reason, in mixed classes with children from multiple grades, only children in grades 1 and 5 were eligible. Median number of children in invited classes was 20 (range 6-27, interquartile range 18-22). We aimed to enrol at least three classes and at least 40 children in each school level at the invited schools.

Children and adolescents (hereafter referred to as children) of the selected classes were invited at each testing round, regardless of their participation in the previous round. Major exclusion criterion was suspected or confirmed SARS-CoV-2 infection in the given child during the testing (precluding the attendance of the testing at school). Children dropped out of the study if they moved to a non-participating school.

### Testing timeline

Venous blood samples were collected from participating children at schools in three testing rounds in 2020-2021. In the first round of testing (T1) on June 16 – July 9, 2020, 2601 children participated; in the second testing (T2) on October 26 – November 19, 2020, 2552 children participated; and in the third testing round (T3) in March 15 – April 16, 2021, 2487 children participated. Flowchart of study participants is shown in Figure 1. The count of children for whom serology results were available at different combinations of the three testing rounds is shown in Appendix 2.

### Serological test and outcomes

Venous blood samples were analysed with ABCORA binding assay of the Institute of Medical Virology (IMV) of the University of Zurich, which is based on Luminex technology. The test is described in detail elsewhere [17]. The test analyses immunoglobulins G (IgG), M (IgM) and A (IgA) against four SARS-CoV-2 targets (receptor binding domain (RBD), spike proteins S1 and S2, and the nucleocapsid protein (N), as well as S1 protein of human coronavirus HKU1 (HCoV-HKU1)), yielding 15 different parameters.

The primary serological outcome used in this study was the binary result of ABCORA 2.3 algorithm, which integrates the 15 parameters of the ABCORA test holistically in comparison to the reference pre-pandemic and confirmed infection plasma using random forests approach [17]. The approach allows higher accuracy than the ABCORA 2.0 algorithm (based on individual thresholds for 12 serological parameters), which we used previously in our study [7,13]. Accuracy parameters of ABCORA 2.3 were 98.2% sensitivity and 99.4% specificity (the latter established against children pre-pandemic plasma values). Serological results were unavailable if venous blood could not be obtained or previously enrolled children declined venipuncture during testing.

Serological outcomes of the three testing rounds were combined to construct a composite (cumulative) outcome – proportion of children who tested seropositive in at least one of the testing rounds (ever-seropositive children). The composite outcome allowed to estimate the proportion of all children who had SARS-CoV-2 antibodies (i.e., were infected) any time by a specific testing, rather than cross-sectional seroprevalence at a single testing, and was measured for T1, T2 and T3 separately (outcomes T1, T12, and T123). In addition, to examine the clustering of new cases within tested classes, we defined the outcome of newly seropositive children at T3 (seropositive at T3 but not in the previous testing rounds), similar to the approach in the previous publication [7].

Exact definitions of positive and negative T1, T12 and T123 serological outcomes, based on all possible combinations of test results from the three testing rounds, are provided in Appendix 2. For the population-level estimates, serological outcomes were adjusted for test accuracy parameters and study population structure, as described below.

### Questionnaires and other information sources

We collected questionnaires from parents of participating and declining to participate children, class and school level information from teachers and school principals, and official statistics of SARS-CoV-2 incidence, as detected by RT-PCR testing.

Parents of participants filled an online questionnaire upon enrolment at either T1, T2 or T3, depending on the entry point to the study, and subsequent follow-up questionnaires approximately every three months. Questionnaires covered information on sociodemographic and health information of the child and household.

In March-April 2021, parents of non-participating children in the invited classes were asked to complete an anonymous questionnaire on the reasons for non-participation (paper or online). This questionnaire was distributed in 50 out of the 55 participating schools (5 schools did not agree). Detailed methods and results of this survey are described in Appendix 4.

Teachers of the invited classes provided information doe the tested classes on the total number of children and children absent on the day of testing of T3 (March-April 2021) due to diagnosed or suspected SARS-CoV-2 infection, quarantine, or other reasons.

After T3 testing, we performed semi-structured telephone interviews with school principals of schools in which 2 or more classes with clusters of newly T3 seropositive children (defined as 3 or more such children) were detected. Collected information included the numbers of RT-PCR diagnosed and quarantined children and their main teachers in the period between T2 and T3 in the affected classes, temporal sequence of diagnoses and quarantine, and additional information on potential index and secondary cases. One interviewer performed all interviews. Intraclass transmission was deemed possible if time window between any reported RT-PCR diagnosed or quarantined children was within 2 weeks and unlikely if longer. We did not assess intraclass transmission if no confirmed or suspected SARS-CoV-2 infection among class children was reported.

We obtained official statistics of RT-PCR confirmed SARS-CoV-2 infections in the canton of Zurich [18] in order to calculate cumulative incidence of diagnosed SARS-CoV-2 cases by T1, T2 and T3 testing, in children aged 7-17 years. Cumulative incidence was calculated from the first SARS-CoV-2 case in Zurich on February 2 to June 30, 2020 (median time point of T1), to November 6, 2020 (T2), and to March 29, 2021 (T3).

### Statistical analysis

Descriptive statistics included summaries of participation rates at the school, class and individual level at each testing round. We report key characteristics of participants (age, school level, sex) as well as their reported symptoms summarized as median (range) or count (%). We did not include children without serology results or without filled questionnaires on symptoms in the analysis of the relevant outcomes. There were no missing data of age, sex, class and school identity.

We calculated the participation rate in a class as the ratio of tested children to all invited children in the class. Proportion of absent children on the day of testing at T3 was calculated to the total number of children in the classes. We calculated the participation rate in the survey of non-participants to the total of non-participating children in the invited classes who were present at school at the day of testing (and thus received the survey).

To estimate the proportion of ever-seropositive children by T1, T2 and T3 testing, we employed Bayesian logistic regression [19], which adjusted for participants’ grade at school, sex and geographic district of the school, and contained random effects for school levels (lower, middle and upper). The Bayesian approach permitted adjusting for the sensitivity and specificity of the SARS-CoV-2 antibody test and the hierarchical structure of cohort (individual, school and district levels). To compute estimates representative for the canton of Zurich, we post-stratified our results according to the total population size at the school level and geographic district. We compared the estimates at T3 for the lower, middle and upper school levels for statistically significant differences.

Masks were introduced for upper school level children in October 2020. Masks were also obligatory for middle school level children from late January 2021; however, the majority of SARS-CoV-2 infections between T2 and T3 happened earlier, in November 2020 – January 2021. We employed difference-in-differences (DD) analysis [20] to examine whether upper school level had different than expected seroprevalence at T3, in regard to the previous trend at T1-T2 and in comparison to middle and lower school levels. Briefly, assuming that the outcome (seroprevalence) would develop in parallel over time in the compared groups (different school levels), DD allows identifying potential deviation from the parallel trend in the intervention group. We modelled DD and examined the parallel trends assumption with a linear probability model. The model included children who were tested three times (n=1965). The binary outcome was ever-testing-positive by T1, T2 and T3. The models controlled for the measurement time point (T1, T2 or T3) and class of the child (thus, implicitly also for district and school grade). Robust cluster-corrected errors at the child level were used. We applied the model to compare upper school level with lower, middle, and combined lower and middle school levels.

We compared the cumulative incidence of RT-PCR confirmed SARS-CoV-2 cases with the proportion of ever-seropositive children, in order to estimate the ratio of undiagnosed to all SARS-CoV-2 cases.

### Analysis of potential clusters

We further examined the potential transmission within classes with at least 3 newly T3 seropositive children (potential cluster). The threshold of 3 or more children was chosen as over a period of 5 months with high community transmission between T2 and T3 testing rounds, two seropositive children in a class are very likely to be detected even if infections are not associated. Thus, three or more cases indicate that potentially transmission could have occurred within a class – even though the associations and temporal sequence cannot be determined with serological testing. Potential clustering was examined in a subset of classes with high participation, in which at least 5 children and at least 50% of the children were successfully tested at the relevant time point. This way, classes with low participation rate were excluded as potentially underpowered to detect potential clusters.

We compared the observed distribution of seropositive children within the classes to the distribution expected if SARS-CoV-2 infections were distributed among children across classes independently (randomly). The approach is described in detail in a previous publication [7]. In the simulation study, we created 2500 hypothetical populations of children, corresponding to the study population in terms of the observed overall seroprevalence, number of classes and tested children within them. Only classes with high participation rate were considered, and only newly T3 seropositive or T3 seronegative children (thus, excluding those who tested positive in the previous rounds). We further ran the same simulation study for classes within the lower, middle and upper school levels separately. The outcome of interest in the simulation study was the expected distribution of classes with 3 or more newly T3 seropositive children. By comparing the expected and the observed distributions, we could estimate whether the observed number of classes with clusters is compatible with the hypothesis of no association of SARS-CoV-2 infections within classes. We further compared the results of the simulation to the information obtained in school principal interviews, whether intra-class transmission was possible.

Data analysis was performed with R version 4.0.3 [21]. Bayesian hierarchical modelling was performed using the R package rstan [22].

### Patient and public involvement

As described in the previous publications [7], several school principals were consulted during the development of the protocol to ensure feasibility of the planned study. Feedback was collected from invited and enrolled children and parents, and school principals, in order to adapt the communication strategy and channels, and improve the implementation of the study. Online informational sessions encouraging open exchange and feedback were organised at the onset of the study for invited and enrolled school principals, personnel and parents of the children. Participants could further share feedback via dedicated phone number (hotline) and email. Results of individual tests were communicated to the participants, and individual school-level results disseminated to participating schools.

Findings were disseminated in lay language in the national and local press, to the national and regional educational and public health departments and on the website of the study (www.ciao-corona.ch).

## Results

In total, 2487 children from 275 classes in 55 schools in the canton of Zurich were tested in March 15 – April 16, 2021 (T3). Of these, 2237 (90%) participated in the T2 testing in October-November 2020, and 2176 (87%) in the T1 testing in June-July 2020. Retention rate from T1 to T2 was 84% (2176/2601) from T1 to T3, and 88% (2237/2552) from T2 to T3.

Serological results were available for 2450/2487 (99%) of participating children at T3. The flowchart of children enrolled and tested at each of the three testing rounds is shown in Figure 1.

Serological results were available at T3 for 768 children in lower school level (median age 8, age range 7-10 years), 845 children in middle school level (median age 12, age range 8-13 years) and 837 children in upper school level (median age 14, age range 12-17 years). From them, 1279 children reported female, 1165 male, and 6 other gender. The median participation rate within invited classes was 50% (interquartile range 35% - 63%; median was 45% in lower, 54% in middle and 50% in upper school levels).

### Serological results

Table 1 presents the distribution of serological results at T1-T3 of children who were tested in all three testing rounds (see Appendix 2 for distribution of results of children who were tested in only one or two rounds). Among children with serological results in all three testing rounds, from 44 testing positive at T1, 29 tested positive again at T2, and from these 26 also in T3 (Table 1). Among children with serological results in the respective two rounds of testing, 33/52 (63%) of those seropositive at T1 were seropositive after 4 months at T2, 32/46 (70%) after 9 months at T3, and 101/115 (88%) of those seropositive at T2 were seropositive after 5 months at T3 (Appendix 2).

**Table 1.** Longitudinal serological results of children tested at all three testing rounds (n=1965)

| T1 result |  | T2 result |  | T3 result |  |
| --- | --- | --- | --- | --- | --- |
| Positive | 44 | Positive | 29 | Positive | 26 |
|  |  |  |  | Negative | 3 |
|  |  | Negative | 15 | Positive | 5 |
|  |  |  |  | Negative | 10 |
| Negative | 1921 | Positive | 73 | Positive | 65 |
|  |  |  |  | Negative | 8 |
|  |  | Negative | 1848 | Positive | 209 |
|  |  |  |  | Negative | 1639 |

T1 – June 16-July 9, 2020; T2 – October 26-November 19, 2020; T3 – March 15-April 16, 2021.
The total number of serological results at each of T1, T2, and T3 is 1965. The table depicts all possible combinations of test results at the three testing rounds. For example, from 44 children who tested positive at T1, 29 tested positive at T2 (from them – 26 tested positive and 3 negative at T3) and 15 tested negative at T2 (from them – 5 positive and 10 negative at T3).

Figure 2 shows the proportion of ever seropositive children by T1, T2 and T3 in the total study population, by school levels and in districts of the canton of Zurich. The proportion was 1.5% (95% credible interval (CrI) 0.6% to 2.6%) at T1, 6.6% (95% CrI 4.0% to 8.9%) at T2 and 16.4% (95% CrI 12.1% to 19.5%) at T3.

**Figure 2.**
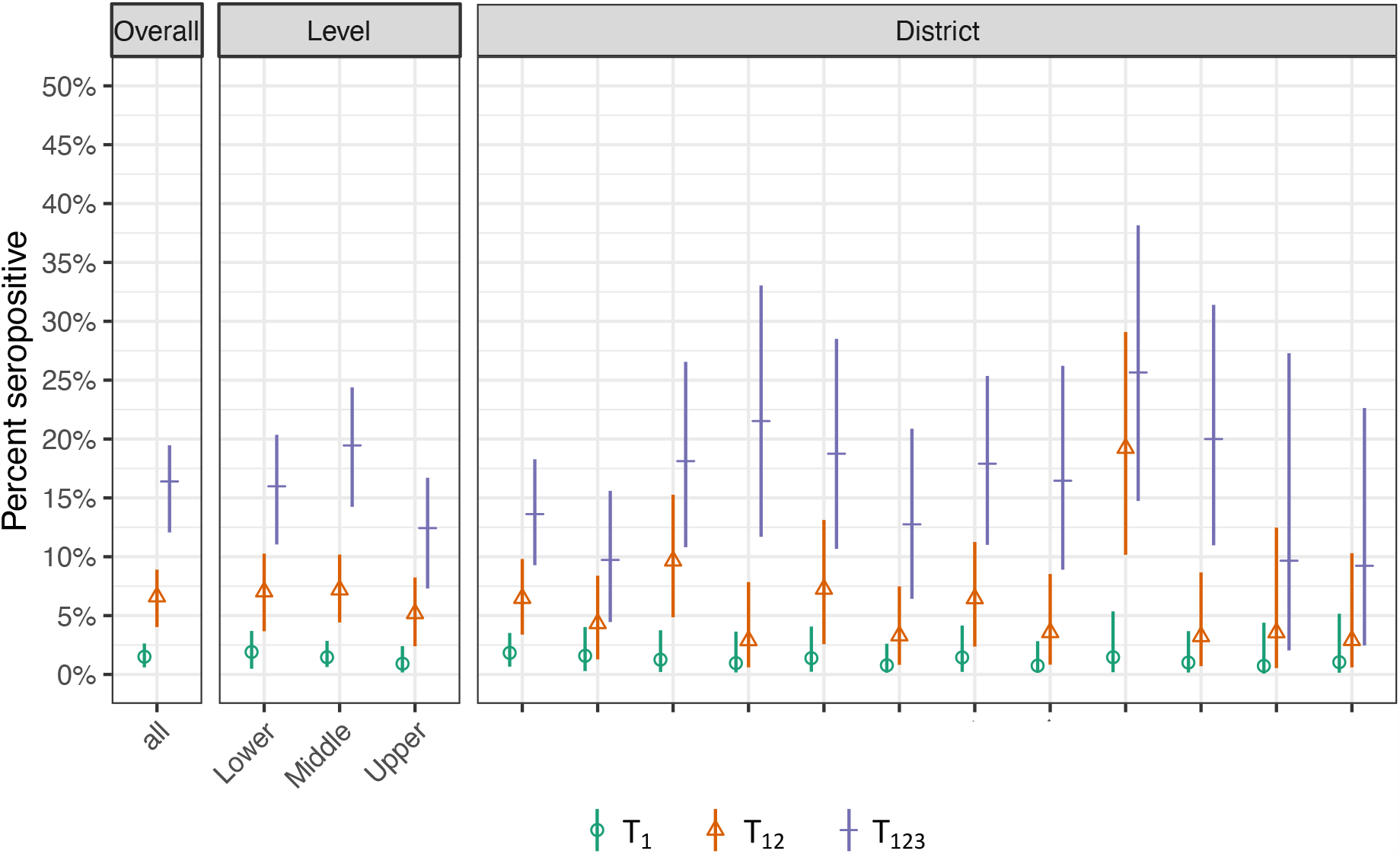
Proportion of ever seropositive children in June-July 2020 (T_1_), October-November 2020 (T_12_) and March-April 2021 (T_123_) T_1_ – proportion of children testing seropositive by June-July 2020, T_12_ – by October-November 2020, T_123_ – by March-April 2021. Districts are ranked by their population size, from largest to smallest. Overall and school level specific estimates (lower school level: grades 1-2, children aged 6-10 years, middle school level: grades 4-5, children aged 9-13 years, upper school level: grades 7-8, children aged 12-16 years; grades and age range are reported for time point of T1), and district specific estimates for the canton of Zurich, Switzerland. Districts are ranked in the order of decreasing population size.

Proportion of ever seropositive children at T3 was 16.0% (11.0% to 20.4%) in lower, 19.5% (14.2% to 24.4%) in middle, and 12.4% (7.3% to 16.7%) in upper school levels. Seroprevalence was not statistically significantly different between lower and middle levels (p=0.26) or lower and upper levels (p=0.18), but different between middle and upper school levels (p=0.02). The proportion of ever seropositive children ranged from 9.2% to 25.7% in the geographic districts of Zurich at T3. The proportion did not differ between boys and girls at any of the testing rounds, although the estimates were slightly higher for boys (T1: 1.8% vs 1.1%; T2: 7.1% vs 6.1%; T3: 17.2% vs 15.6%).

In the difference-in-differences analysis, parallel trends assumption between T1 and T2 was valid based on visual inspection (Appendix 3A) and upon formal examination with the linear probability models (Appendix 3B). In comparison to lower and middle school levels, the proportion of ever-seropositive children in the upper school level was lower at T3 than expected (absolute difference of 3.2% (95% CI -7.0% to 0.6%, p=0.098), relative difference of 19%). The proportion was lower by 5.1% (95% CI -9.4% to -0.7%, p=0.022, relative difference of 27%) when comparing only to the middle school level, and lower by 0.8% (95% CI -5.4% to 3.8%, p=0.718, relative difference of 6%) when comparing only to the lower school level.

The ratio of children testing positive for SARS-CoV-2 infection with RT-PCR test to those testing ever-seropositive was 1 to 21.7 by June-July 2020 (T1), 1 to 5.8 by October-November 2020 (T2), and 1 to 3.5 by March-April 2021 (T3).

### Symptoms

Symptoms between T2 and T3 were reported in 67/182 (37%) of newly seropositive and 235/1443 (16%) of seronegative children (Figure 3). The most commonly reported symptoms among children who were newly seropositive were fatigue (30/182, 17%), sore throat (30/182, 17%), headache (27/182, 15%), fever (26/182, 14% for fever ≥38°C and 25/182, 14% for subjective fever) and runny or congested nose (22/182, 12%). Fatigue, sore throat, headache, fever, stomach ache, muscle or joint pain, and loss of smell or taste were reported more frequently by seropositive than seronegative children.

**Figure 3.**
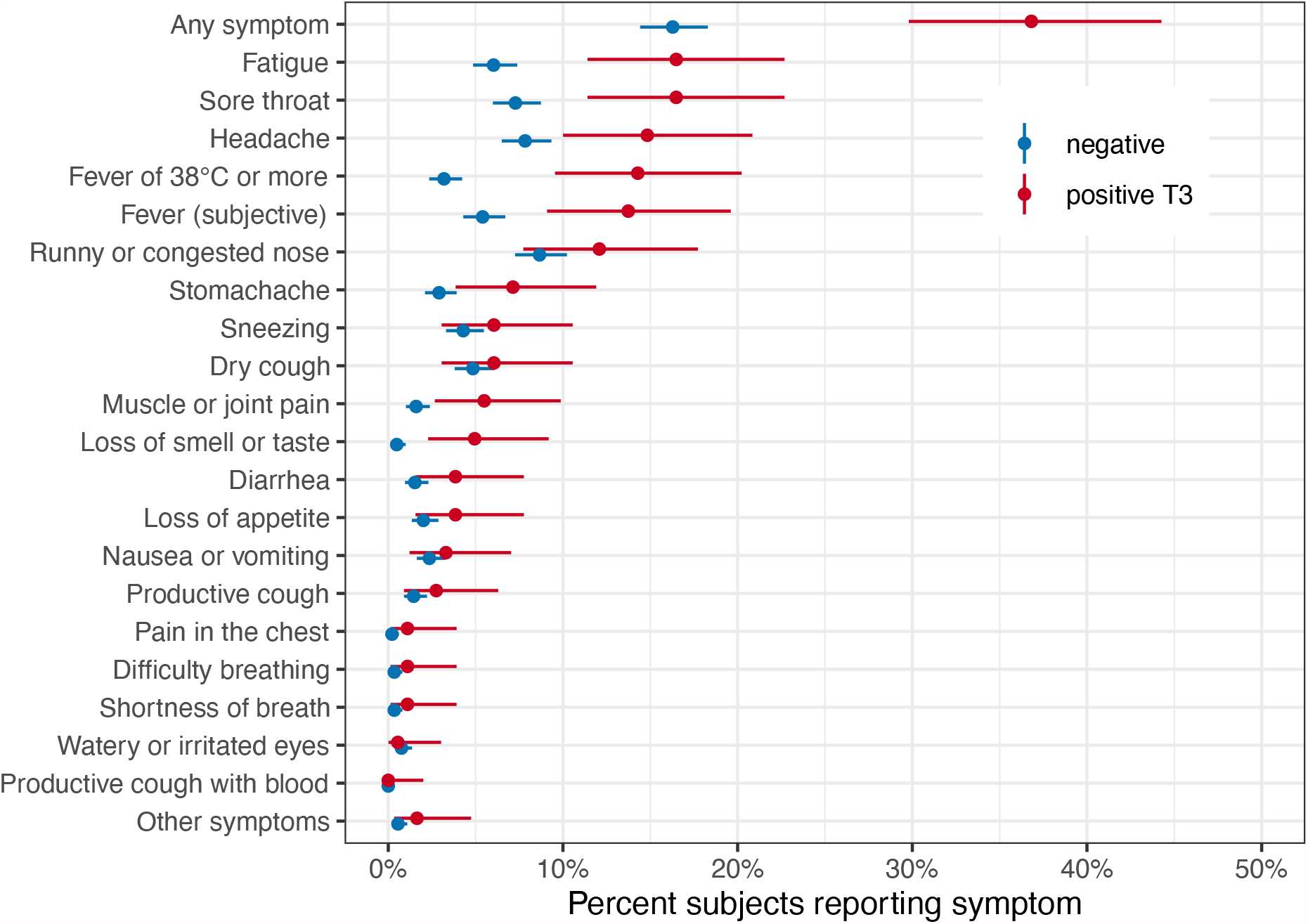
Symptoms reported between October-November 2020 and March-April 2021 in children who were seronegative and newly seropositive at T3 (March-April 2021)

Similar proportion of seropositive children reported symptoms in the different school levels: 35% (22/63) in lower, 40% (32/81) in middle, and 37% (14/38) in upper school levels. However, among the seronegative, older children were less likely to report symptoms (21% (101/484) in lower, 16% (78/493) in middle, and 13% (60/466) in upper school levels).

Symptoms among the newly seropositive children were reported the most frequently in December 2020 and January 2021. None of the participating children reported hospitalisation between T2 and T3 testing.

### Cluster analysis

At least one child who was *newly* seropositive at T3 was detected in 53 of 55 schools and 151 of 275 (55%) classes (76 of 119 (64%) classes with high participation rate). At least one child who was *ever* seropositive at T3 was detected in all 55 schools and 184 of 275 (67%) classes (95 of 119 (80%) classes with high participation rate). The number of children who were *newly* seropositive at T3 ranged from 0 to 13 in a school level (two adjacent grades), and from 0 to 7 in a class. The number of children who were *ever* seropositive at T3 ranged from 0 to 15 within a school level, and from 0 to 13 within a class. Figure 4 shows the distribution of children who were seropositive by T1, T2 and T3 within the 119 classes with high participation rate at T3.

**Figure 4.**
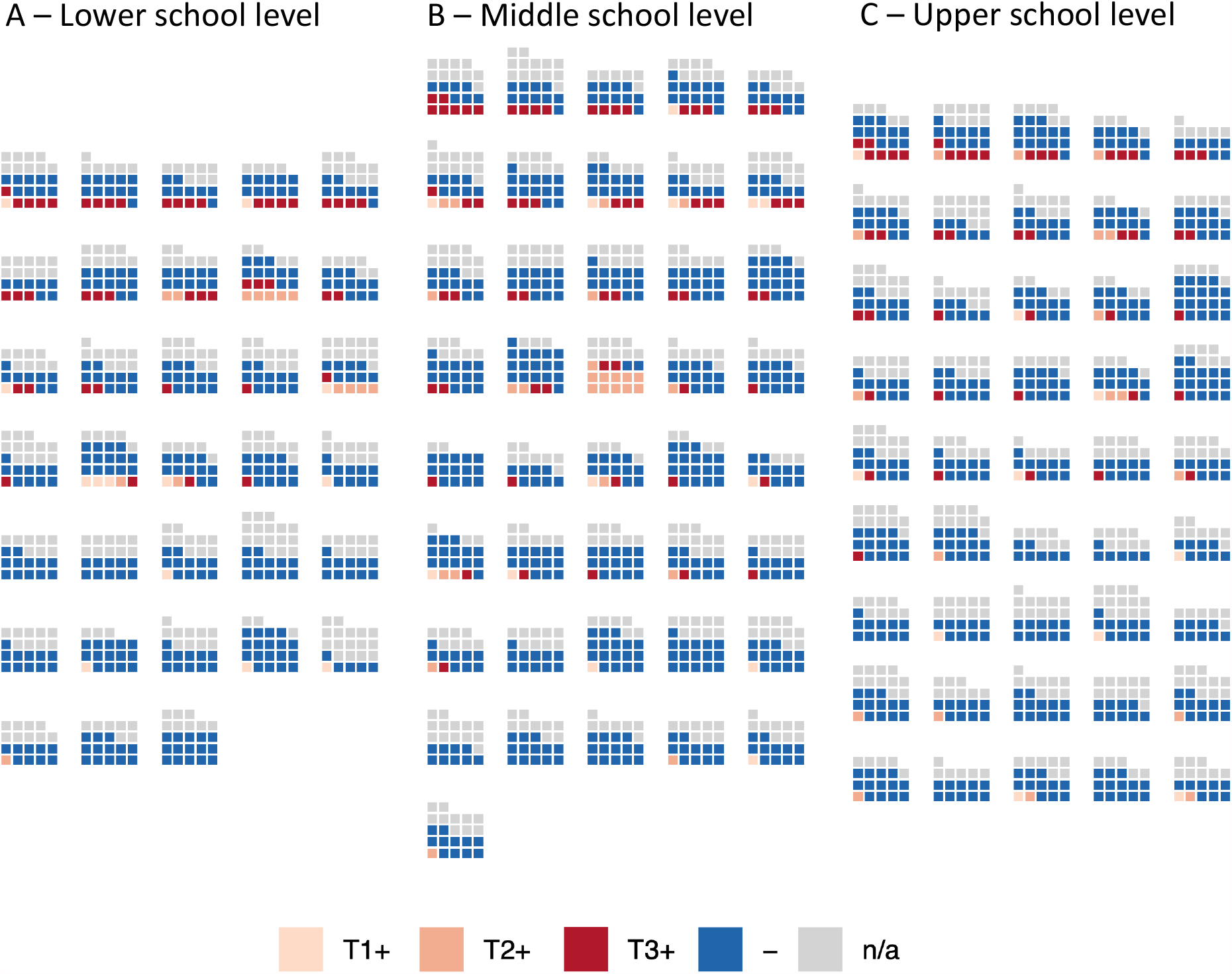
Distribution of children who were seropositive at T1, T2 and T3 in classes with high participation rate at T3. Each block of squares represents a class. High participation rate means that at least 5 and at least 50% children were tested in the class. T1+ (light pink) – children who tested seropositive at T1 (June-July 2020), T2+ (middle orange) – children who tested seropositive at T2 (October-November 2020) but not T1, T3+ (dark red) – children who tested seropositive at T3 (March-April 2021) but not at T1 or T2, - (dark blue) – children who tested seronegative at T3 and did not test seropositive previously, n/a (grey) – children without serological result at T3.

In total, 39 of 275 (14%) classes had 3 or more children who were newly seropositive at T3: 13 of 94 (14%) in lower school level, 19 of 83 (23%) in middle, and 7 of 98 (7%) in upper school levels. 24 of 119 (20%) classes with high participation had such clusters: 9 of 33 (27%) in lower school level, 10 of 41 (24%) in middle, and 5 of 45 (11%) in upper school level. Classes with clusters were present in 24 schools (14 schools with one, 6 schools with two, 3 schools with 3, and one school with 4 classes with clusters).

From 25 classes with potential clusters, on which information was collected during the interviews with school principals, based on the timing of RT-PCR diagnosis and quarantine in children, teachers and household members of children, intra-class transmission could have happened in 12 classes (63%), was improbable in 7 (37%), and could not be assessed due to insufficient information in 6 classes (Appendix 4). A total of 6 teachers and 40 children were reported as testing RT-PCR positive between T2 and T3, compared to 84 children testing newly seropositive at T3 in these classes. 5 teachers and 55 children were quarantined between T2 and T3, in addition to two whole classes and one whole school. The majority of RT-PCR diagnosed infections and exposure resulting in quarantine in children originated from children’s households. Further details on interview results are summarized in Appendix 4.

Assuming a uniform 10.9% rate of newly seropositive children among those not previously tested positive (158/1422 within the classes with high participation), we would expect to observe a median of 17 (95% CrI 12 to 22) classes with 3 or more seropositive children out of the 119 classes with high participation. In comparison, we observed 24/119 (20%) such classes, making the hypothesis of independent distribution of all seropositive children within classes unlikely (p=0.0052). The simulated distribution of expected number of classes with clusters, standardized to 100 such classes, is depicted in Figure 5. In lower school level, a median of 5/33 such classes with clusters was expected based on simulation results compared to 9/33 observed (p=<0.001), in middle school level – 9/41 expected compared to 10/41 observed (p=0.27), and upper school level – 4/45 expected compared to 5/45 observed (p=0.29).

**Figure 5.**
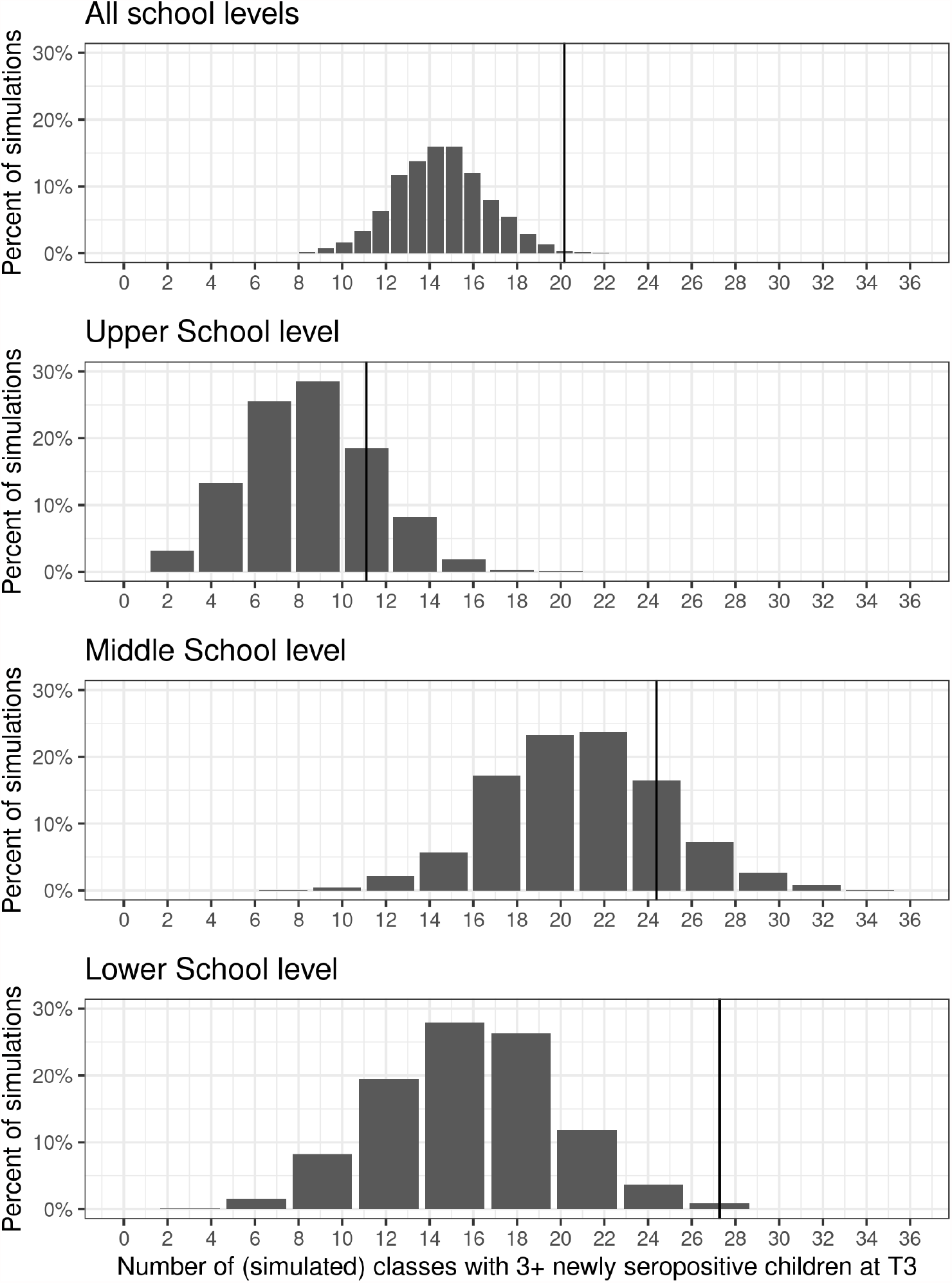
Distribution of expected number of classes with clusters (3 or more newly seropositive children), standardised to 100 classes with high participation rate. Vertical line signifies the number of clusters observed in this study, as standardised to 100 classes (all school levels – 20 (corresponding to 24/119), upper school level 11 (5/45), middle level 24 (10/41), and lower school level 27 (9/33)).

### Reasons for participation and non-participation

At the day of testing at T3, 10 of 5517 (0.2%) children in the invited classes were not at school due to diagnosed or suspected SARS-CoV-2 infection, 47 (0.9%) due to quarantine, and 164 (3.0%) for other reasons (including non-SARS-CoV-2 illness). Parents of 712 of 2659 (27%) eligible but non-participating children provided responses to the survey on reasons for non-participation. From those who reported having received information or invitation to the study, the most common reason for non-participation was the child’s fear of the blood sampling (358/564, 64% of children; 189/276, 69% of girls and 158/259, 61% of boys). Fear of blood sampling was reported more frequently for children in lower school levels (191/264, 72%) than in middle (85/153, 56%) and upper school levels (79/141, 56%) (Figure 6). Based on free-text comments provided by parents, 78% (266/341) of children did not participate due to reasons that could not be modified by the study team (non-modifiable). Among the participants, 28% (374/1331) reported participating for personal reasons (such as for a personal serological test result), and 90% (1192/1331) due to societal reasons (such as contribution to research or containment of the pandemic). Detailed results of these surveys are provided in Appendix 5.

**Figure 6.**
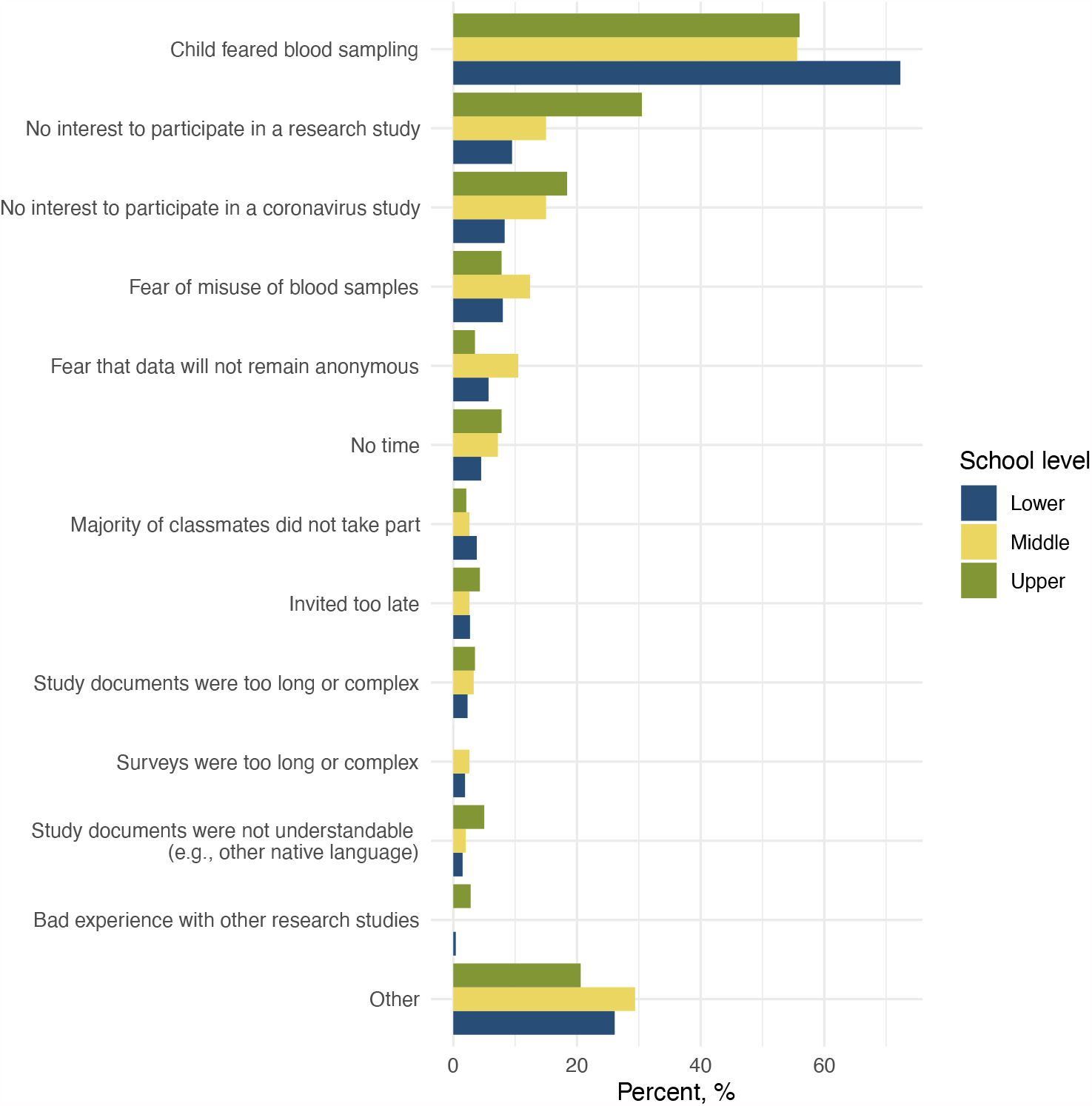
Frequency of reported reasons of non-participation in lower, middle and upper school levels. Lower school level: grades 2-3, children aged 7-10 years, middle school level: grades 5-6, children aged 8-13 years, upper school level: grades 8-9, children aged 13-17 years.

## Discussion

In this cohort study of more than 2500 children in 55 schools tested three times over a period of 10 months, the proportion of children who tested seropositive for SARS-CoV-2 increased from 1.5% in June-July 2020 to 6.6% in October-November and to 16.4% in March-April 2021. In contrast to the previous testing rounds, seroprevalence was lower in upper school level (12.4%, ages 10-17) in comparison to middle (19.5%) and lower (16.0%) school levels. Although potential clusters of at least 3 newly seropositive children were detected in approximately 20% of classes, the majority of them could be explained by SARS-CoV-2 infections not associated within a class, particularly in the middle and upper school levels. In contrast to previous testing rounds in summer and autumn of 2020, newly seropositive children reported a history of acute symptoms more frequently than seronegative children, although 63% of seropositive children still did not report any symptoms. 88% of seropositive children retained their antibodies for at least 5 months, between October 2020 and March 2021.

The canton of Zurich experienced an early and stark second wave of SARS-CoV-2 pandemic in 2020. Daily incidence of RT-PCR cases peaked at 88 in 100’000 inhabitants on October 28 [23], comparable to the peaks of 75 and 90 new cases in 100’000 inhabitants in the US and UK, respectively, in early January 2021 [1]. In parallel to the general population, the number of new cases was rising also in children and adolescents. Physical attendance of schools was not interrupted but mitigation measures were implemented – such as quarantining of symptomatic children, distancing, non-mixing of classes, a school-based contact tracing system, and a gradual adoption of masks for adults and upper and middle school level children. Our study suggests that a significantly higher proportion of children were tested and diagnosed with RT-PCR in late 2020 and 2021 than in the first wave of the pandemic in early 2020: the ratio of seropositive to RT-PCR detected cases in children decreased from 21.7 by June-July 2020 to 3.5 by March-April 2021. This likely reflects the revised indications for RT-PCR testing in children. Initially restrictive, the indications for children over 12 years were changed in September 2020 to match those for adults, and subsequently extended to children over 6 years from March 2021 [24].

The increase in the proportion of seropositive children by March-April 2021 was the smallest (and potential clusters of newly seropositive children less frequent) in upper school level. Age is in general associated with more severe symptoms and worse health outcomes in adults. This higher susceptibility to infection [25,26] is likely related to both biological and behavioural factors. In contrast to our findings, SARS-CoV-2 has been observed to spread more among older children and adolescents [27], partly explained by a different patterns of contacts [28,29]. In Switzerland, adolescents from grade 7 (approximately age 14) were required to wear masks at schools since November 2020. Potentially, consistent masking during the second wave of SARS-CoV-2 infections in November 2020 – January 2021, as well as a different distancing behaviour of teenagers towards their parents compared to young children (given that the majority of infections in children were linked to households), could have contributed to less infections. The preventive effect of masking is supported by the results of the difference-in-differences analysis. In comparison to the middle school level, where masks were introduced three months later, the proportion of children ever testing seropositive in the upper school levels was lower by 5.1% (relative reduction of 27%) than could be expected if seroprevalence developed in the same trend as in the middle school level between T2 and T3. To increase complexity of understanding these age-related differences, no significant difference was found in the development of seroprevalence between T2 and T3 between upper and lower school levels.

Our results from the three testing rounds consistently suggest that only a small part of potential clusters of seropositive children within classes were likely associated with intra-class transmission. Although it is very unlikely that all potential clusters could be explained by chance association of infections, 17 of the affected 24 classes with such clusters (from 119 classes with high participation rate at T3) could be expected by chance, given the overall seroprevalence observed in this study. Therefore, it is also unlikely that more than half of the potential clusters are due to potential spread of the infection within the class or through a common index case. In middle and upper school levels, the observed number of classes with clusters was not significantly different from that expected due to chance association – in contrast to lower school level, where masks for children were not mandated at any time point. This finding was supported by interviews with school principals, in which we found that at least 33% of the clusters were unlikely to be due to intraclass transmission due to more than 2 weeks between confirmed or suspected infections within a class.

In contrast to previous measurements in summer [13] and autumn of 2020 [7], symptoms were reported more frequently in seropositive than seronegative children. It is possible that the retrospective recall of symptoms was influenced by the knowledge of the symptoms, increased attention by participation in previous rounds, population incidence, and personal diagnosis of SARS-CoV-2. The influence of VOC on the symptoms of SARS-CoV-2 infection in children cannot be ruled out. However, the majority of symptom episodes were reported in December 2020 – February 2021 and VOC became predominant in Switzerland only from March 2021 (see Appendix 1) and thus is unlikely to have caused a major proportion of infections in the newly seropositive children. The difference could also be potentially explained as symptoms were assessed over different time periods (June-October 2020 between T1 and T2, October-March 2021 between T2 and T3) at different seasons.

In parallel to testing the participating children, we administered a survey to discover the reasons for non-participation (Figure 6). Detailed reasons for non-participation are rarely known in children cohort studies [30]. Although 50% participation rate in our study is rather high for a study with venous blood sampling in children [31], it could still result in selection bias. Indeed, the predominant reason for non-participation was fear of blood sampling, followed by lack of interest to participate in a research study. Non-participation rate was higher than attrition rate (50% vs 16%), meaning that once enrolled, few children dropped out.

Most of the studies of SARS-CoV-2 spread in schools rely on contact tracing data [3,4,11]. At the start of the 2020/2021 school year, such studies, also supported by a large online survey in the US [32], pointed to the spread being low in schools with implemented protective measures. The few available seroprevalence studies of school children also showed concordant results [7,33,34]. Studies of RT-PCR screening in schools and general population show that they tend to correlate [5,10,35].

Less evidence exists about how SARS-CoV-2 spread within schools might change as VOC with higher infectiousness become more prevalent. The trends are difficult to interpret as a significant proportion of people in high income countries are becoming vaccinated since the first half of 2021. These two important determinants of SARS-CoV-2 infection rate develop independently in different patterns, but also interact. Depending on the transmissibility of the various VOC in the future and their selective transmission among children, relative increase of SARS-CoV-2 infections among children may or may not occur, if schools remain open while other age groups retain reduced contacts or become immunised.

## Strengths and limitations

The *Ciao Corona* cohort study is a population-based cohort of children within randomly selected schools and classes, with repeated measurements of SARS-CoV-2 serology and high retention rate. Ascertainment of SARS-CoV-2 infections via serological testing means that also the asymptomatic, previously not diagnosed children are detected. This is particularly important to provide a complete picture of SARS-CoV-2 infections in children, as under-detection of SARS-CoV-2 infections remains significant.

The study has limitations. First, relying on serological results means that the timing of the infection cannot be known beyond a broad interval between the testing rounds. This means that infection chains cannot be reconstructed as in prospective contact tracing studies, and associations of infections within a class needed to be examined indirectly (e.g., by comparing to the number of clusters expected if all infections were independent) or retrospectively (e.g., by information gathered from school principals). However, these approaches are either probabilistic only, or rely on not always complete or detailed information from school principals. Second, although we used a serological test with high sensitivity and specificity parameters (the latter even specific to the children population), and further adjusted for them in the Bayesian models, false positive and negative results cannot be completely avoided on the individual level. For example, 10-15 children of 393 who tested positive at T3 round of testing could be expected to be false positive, and 5-10 of 2061 who tested negative – false negative. Particularly false positive results could impact the analysis of clustering of positive children (by falsely increasing the number of observed clusters), antibody retention rate (by falsely decreasing the retention), and the difference-in-differences models (where individual children results were used as outcomes). Low prevalence in the initial testing rounds meant that positive predictive value of the serological test was not high (approximately 0.75 at T1 and 0.90 at T2), however, it reached 0.97 at T3. Retention was slightly lower between T1 and T2 than between T2 and T3 potentially as more false positives were expected among T1 participants. Third, although participation rates were high for a study involving venous blood sampling in children, selection bias cannot be ruled out. However, the majority of non-participating parents reported that non-participation was driven by their child’s decision and fear of venous blood sampling – reasons not likely associated with particularly biased selection into the study. Yet, participation bias within the non-participating parents in this survey is still possible.

## Conclusion

Since the beginning of the pandemic in Switzerland in 2020, an increase of SARS-CoV-2 seroprevalence from 1.5% in June-July 2020 to 16.4% in March-April 2020 was accompanied by an increase of potential clusters of seropositive children in classes. Despite schools remaining open since May 2020, the majority of the detected potential clusters in classes could be explained by unrelated, independent infections, likely stemming from household or community transmission. The increase in seroprevalence and clustering was lower in upper school level, where masks were introduced in November 2020, compared to lower and middle school levels. Preventive measures including contact tracing system in schools and improved detection of infections in children possibly contributed to mitigate the spread of SARS-CoV-2 within schools.

## Data Availability

Data is still being collected for the cohort study Ciao Corona. Deidentified participant data might be available on reasonable request by email to the corresponding author at later stages of the study.

## Author’s Contributions

SK and MAP initiated the project and preliminary design, with support of JF. SK, MAP, CB, TR and AU developed the design and methodology. SK, AU, TR, PA recruited study participants, collected and managed the data. SRH performed statistical analysis. AT and IA developed the serology analysis plan, supervised, conducted and evaluated the serology tests. AU wrote the first draft of the manuscript. All authors contributed to the design of the study and interpretation of its results, and revised and approved the manuscript for intellectual content. SK is the guarantor and accepts full responsibility for the work and the conduct of the study, had access to the data, and controlled the decision to publish. The corresponding author SK attests that all listed authors meet authorship criteria and that no others meeting the criteria have been omitted.

## Acknowledgements

We would like to acknowledge Miquel Serra-Burriel (Epidemiology, Biostatistics and Prevention Institute, University of Zurich) for his valuable inputs on the implementation of the difference-in-differences analysis.

## Ethics approval

The study was approved by the Ethics Committee of the Canton of Zurich, Switzerland (2020-01336). All participants provided written informed consent before being enrolled in the study.

## Data sharing statement

Data is still being collected for the cohort study *Ciao Corona*. Deidentified participant data might be available on reasonable request by email to the corresponding author at later stages of the study.

## Transparency declaration

The lead authors affirm that the manuscript is an honest, accurate, and transparent account of the study being reported, no important aspects of the study have been omitted, and any discrepancies from the study as originally planned and registered have been explained.

---

**Appendix 1** Weekly incidence of SARS-CoV-2 detected cases and the proportion of the variants of concern (VOC) among them in September 2020 – April 2021

**Appendix 2** Combinations of longitudinal serological results, at three testing time points, and serological outcomes

**Appendix 3** Difference-in-differences model of the change in ever-seroprevalence between T2 and T3 in upper school level

**Appendix 4** Detailed information of classes with potential clusters (3 or more newly seropositive children at T3) in schools with at 2 or more classes with clusters

**Appendix 5** Detailed results of survey for participants and non-participants on reasons for participation and non-participation

