## Appendices 1-3 for "Evolution of SARS-CoV-2 seroprevalence and clusters in school children from June 2020 to April 2021 reflect community transmission: prospective cohort study *Ciao Corona*"

**Appendix 1** Weekly incidence of SARS-CoV-2 detected cases and the proportion of the variants of concern (VOC) among them in September 2020 – April 2021

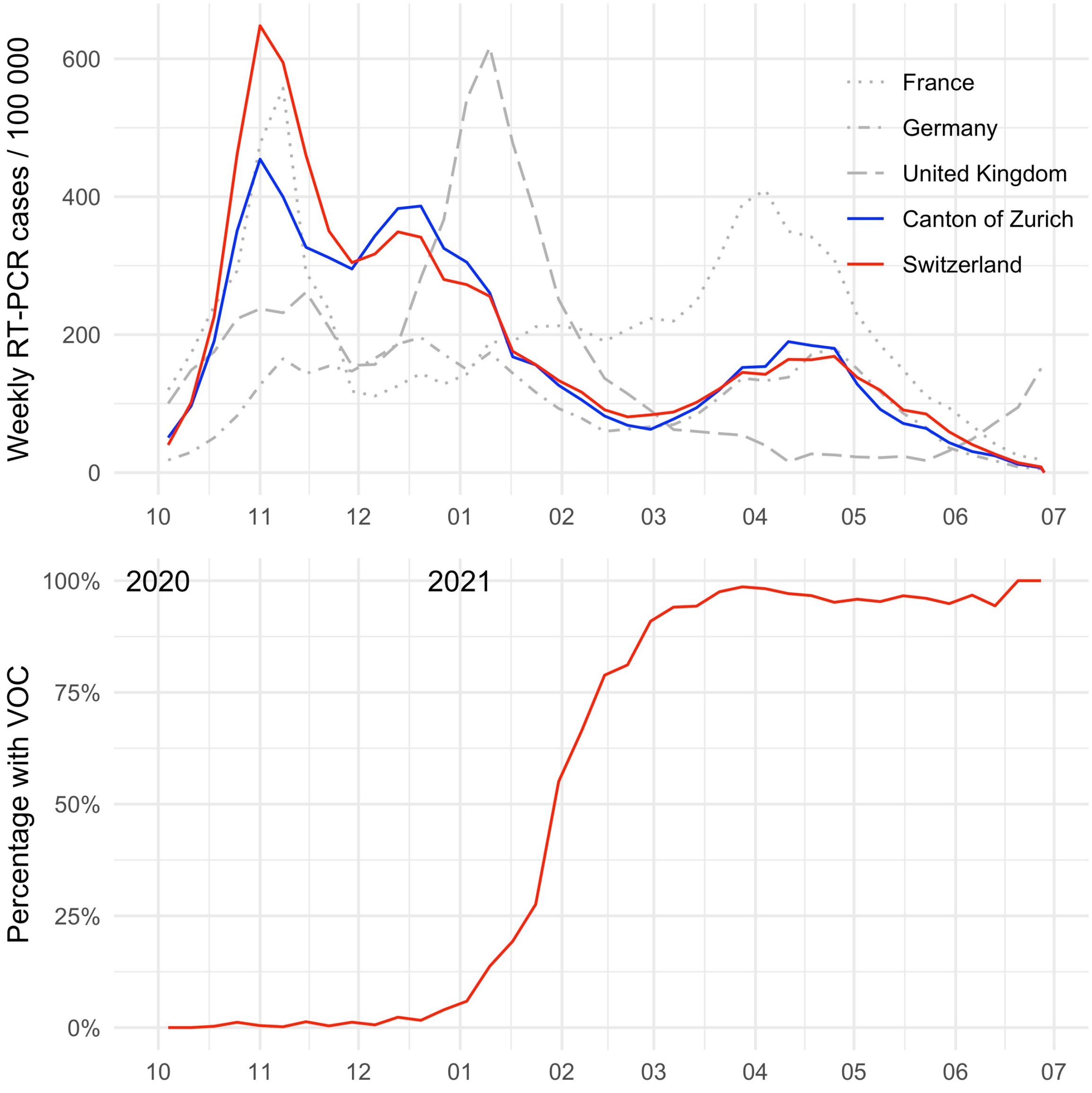


Percentage with VOC was measured for Switzerland, based on a representative sample of approximately 2000 positive samples per week.

Data source: Swiss Federal Office of Public Health, https://opendata.swiss/de/dataset/covid-19-schweiz (accessed July 2, 2021), Our World In Data, https://github.com/owid/covid-19-data/tree/master/public/data (accessed July 15, 2021)

**Appendix 2** Combinations of longitudinal serological results, at three testing time points, and serological outcomes

| Serology test results | | | | Serological outcomes | | |
|---|---|---|---|---|---|---|
| **T1** | **T2** | **T3** | **N** | **$T_1$** | **$T_{12}$** | **$T_{123}$** |
| + | + | + | 26 | + | + | + |
| + | + | - | 3 | + | + | + |
| + | + | NA | 4 | + | + | + |
| + | - | + | 5 | + | + | + |
| + | - | - | 10 | + | + | + |
| + | - | NA | 4 | + | + | + |
| + | NA | + | 1 | + | + | + |
| + | NA | - | 1 | + | + | + |
| + | NA | NA | 2 | + | + | + |
| - | + | + | 65 | - | + | + |
| - | + | - | 8 | - | + | + |
| - | + | NA | 12 | - | + | + |
| NA | + | + | 10 | | + | + |
| NA | + | - | 3 | | + | + |
| NA | + | NA | 7 | | + | + |
| - | - | + | 209 | - | - | + |
| NA | - | + | 22 | | - | + |
| - | NA | + | 27 | - | | + |
| NA | NA | + | 28 | | | + |
| - | - | - | 1639 | - | - | - |
| NA | - | - | 179 | | - | - |
| - | NA | - | 89 | - | | - |
| NA | NA | - | 129 | | | - |
| - | - | NA | 225 | - | - | |
| NA | - | NA | 69 | | - | |
| - | NA | NA | 154 | - | | |
| NA | NA | NA | 47 | | | |
| **Number of positive results (red)** | | | | 56 | 161 | 447 |
| **Total number included in the analysis of this outcome (coloured)** | | | | 2484 | 2504 | 2483 |
| **Raw proportion of seropositive results** | | | | 2.3% | 6.4% | 18.0% |

Results in the coloured cells are included in the analysis of the specific outcome (column). $T_{12}$ and $T_{123}$ outcomes are binary (red – ever-seropositive and blue – seronegative), but the seropositive can be further considered by the time they first tested seropositive (different hues of red for T1, T2, and T3). For $T_{12}$ and $T_{123}$ outcomes all negative results of the current relevant testing and positive results of current and previous testing rounds are included. The colours of the cells correspond to the colours used in Figure 4 of the manuscript (light orange – newly seropositive at T1, middle red – newly seropositive at T2, dark red – newly seropositive at T3, blue – negative).

**Appendix 3** Difference-in-differences model of the change in ever-seroprevalence between T2 and T3 in upper school level

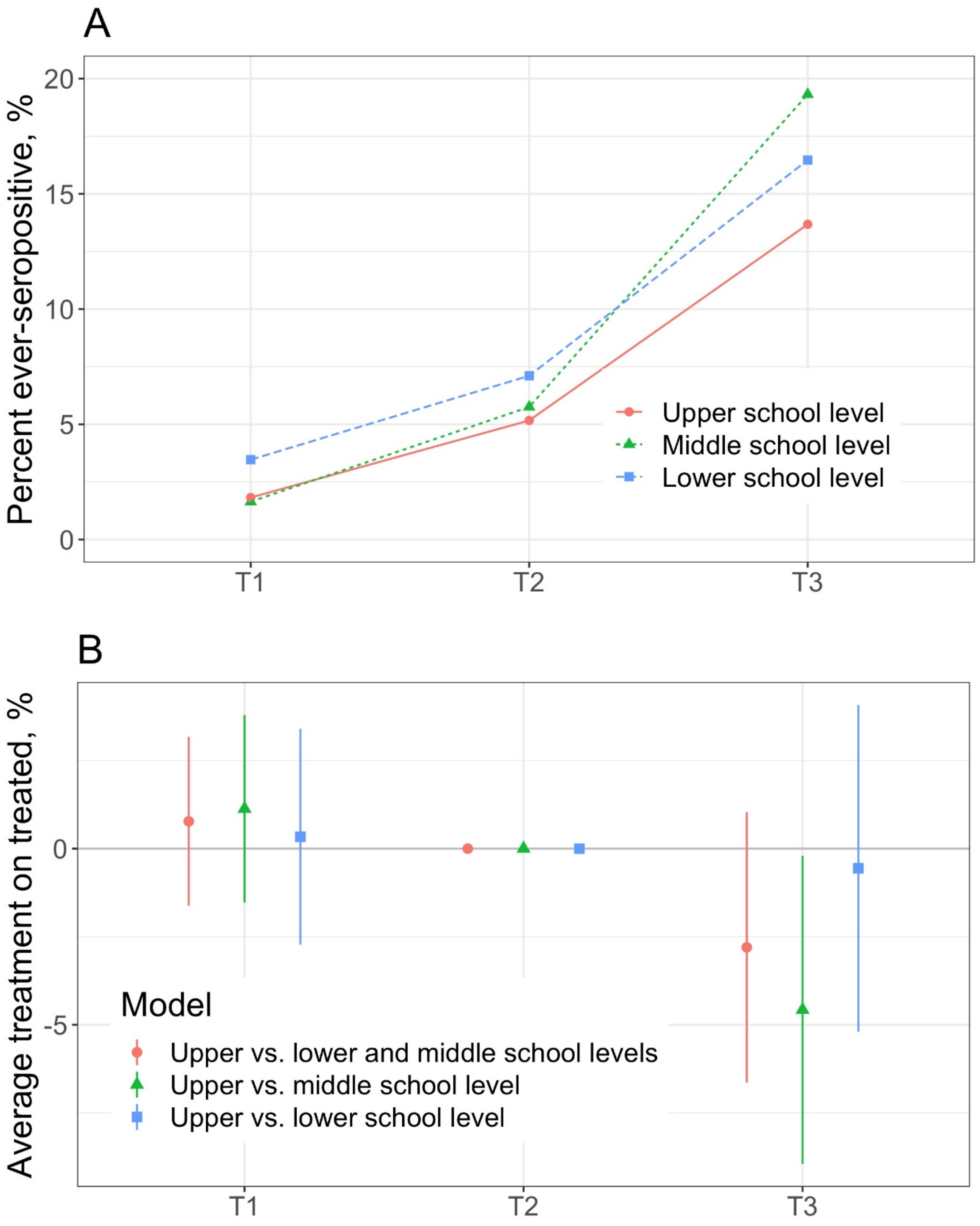


A – Raw proportion of children, included in the models, ever testing seropositive by T1, T2 and T3 time points.

B – Difference-in-differences estimates of linear probability models, with T2 as the reference. Parallel trends assumption is valid, as the estimates at T1 are not different from the T2 reference.
