## Appendix 4 for "Evolution of SARS-CoV-2 seroprevalence and clusters in school children from June 2020 to April 2021 reflect community transmission: prospective cohort study *Ciao Corona*"

**Appendix 4** Detailed information of classes with potential clusters (3 or more newly seropositive children at T3) in schools with 2 or more classes with clusters

| School | School level | N of pupils: T1+/T2+/T3+<br>negative<br>missing* | RT-PCR positive in T2-T3 | | Quarantine/ isolation in T2-T3 | | Information**<br>(*unrelated time points* signify >2 weeks interval between all diagnosed or potential infections) | Possible index cases of individual pupils*** | | | Transmission in class plausible **** | Comments |
|---|---|---|---|---|---|---|---|---|---|---|---|---|
| | | | Teacher | Child | Teacher | Child | | Teacher | Child | House-hold | | |
| 1 | lower | 1/0/4<br>10<br>3 | 0 | 0 | 0 | 3 | 3 pupils in quarantine: n=1 due to PCR+ teacher not related to regular school, n=2 both due to PCR+ household member (HM) of one of the 2 pupils, with whom both had close contact | 0 | 0 | 1 | 1 | |
| | lower | 1/0/3<br>5<br>11 | 1 | 0 | 1 | 7 | 7 pupils in quarantine: n=5 due to PCR+ HM and n=2 for unknown reasons | 0 | 0 | 1 | 1 | |
| | lower | 0/0/4<br>8<br>8 | 0 | 0 | 0 | 4 | 4 pupils in quarantine: n=1 due to PCR+ HM, n=1 due to PCR+ teacher not related to regular school, n=1 due to PCR+ family friend, n=1 for unknown reasons | 0 | 0 | 1 | 0 | |
| | lower | 1/0/4<br>10<br>3 | 0 | 1 | 1 | 4 | 1 pupil in isolation: PCR+ for unknown reasons.<br>3 pupils in quarantine: n=2 each due to PCR+ HM (unrelated time points), n=1 for unknown reason | 0 | 0 | 1 | 0 | |
| 2 | lower | 0/0/4<br>8<br>11 | 0 | 2 | 0 | 3 | 2 pupils in isolation: n=1 first in quarantine due to PCR+ HM then PCR+, n=1 PCR+ due to symptoms.<br>2 pupils in quarantine at unrelated time points each due to PCR+ HM | 0 | 0 | 1 | 1 | Outbreak in grade 5 class non-participating in this study: 8 pupils PCR+, not clear where Index came from, family of 1 pupil and HM of another pupil were tested PCR+ |
| | middle | 0/0/4<br>10<br>6 | 0 | 0 | 0 | 1 | 1 pupil in quarantine due to PCR+ HM | 0 | 0 | 1 | ? | |
| | middle | 0/0/3<br>3<br>16 | 0 | 0 | 0 | 2 | 2 pupils in quarantine at unrelated time points each due to PCR+ HM | 0 | 0 | 1 | 0 | |
| 3 | lower | 0/0/3<br>7<br>9 | 1 | 1 | 1 | whole class | Outbreak in a parallel class due to PCR+ child (probably infected by HM; index HM with severe symptoms but not tested). Then teacher and several children PCR+ in this and the parallel classes, the remaining children not tested; both classes (including this one) quarantined | 0 | 0 | 1 | 1 | Transmission of virus among pupils in the lower school level class probable, primary Index case most probably HM of a child. |

| | | | | | | | | | | | | |
|---|---|---|---|---|---|---|---|---|---|---|---|---|
| | middle | 1/1/3<br>12<br>5 | 0 | 0 | 0 | 1 | 1 pupil in quarantine due to travel abroad after fall vacation | 0 | 0 | 0 | ? | |
| | middle | 1/1/3<br>7<br>8 | 1 | 3 | 1 | 3 | 1 teacher in isolation PCR+, 3 pupils in isolation, each PCR+ due to PCR+ HM (unrelated time points) | ? | 0 | 1 | 1 | |
| 4 | lower | 1/0/3<br>2<br>15 | 1 | 3 | whole school | whole school | 3 pupils in isolation: n=1 PCR+ for unknown reason; n=1 due to PCR+ HM; n=1 PCR+ with symptoms | 0 | ? | 1 | 1 | School outbreak with the whole school in quarantine |
| | middle | 1/1/3<br>7<br>8 | 1 | 7 | | | 1 teacher PCR+, 3-6 days later PCR+ pupil tested because of loss of taste. Testing of the whole class 2 days later - 6 further pupils PCR+, none with symptoms, 5 pupils PCR– | 1 | 0 | 0 | 1 | |
| | middle | 0/1/3<br>7<br>12 | 1 | 6 | | | 6 pupils in isolation: n=1 in quarantine due to PCR+ HM, then tested PCR+ in the outbreak testing together with 1 teacher and 5 further pupils PCR+ | ? | ? | 1 | 1 | |
| 5 | lower | 1/1/3<br>3<br>13 | 0 | 0 | 0 | 0 | No diagnosed or suspected SARS-CoV-2 infections known | ? | ? | ? | ? | |
| | middle | 0/0/4<br>6<br>11 | 0 | 0 | 0 | 0 | No diagnosed or suspected SARS-CoV-2 infections known | ? | ? | ? | ? | |
| 6 | middle | 0/0/3<br>13<br>8 | 0 | 8 | 1 | whole class | First a few children PCR+, then in total 8 PCR+ shortly afterwards. Subsequently class in quarantine, teacher then tested PCR– but later tested seropositive | 0 | 1 | ? | 1 | |
| | middle | 1/1/3<br>3<br>15 | 0 | 3 | 0 | 3 | 3 pupils in quarantine at unrelated time points each due to PCR+ HM | 0 | 0 | 1 | 0 | |
| 7 | upper | 0/1/3<br>10<br>3 | 0 | 2 | 0 | 2 | 2 pupils in isolation at unrelated time points for unknown reasons | 0 | ? | ? | 0 | A class not participating in this study in quarantine during Christmas break due to several PCR+ pupils |
| | upper | 0/0/3<br>7<br>5 | 0 | 1 | 0 | 1 | 1 pupil in isolation for unknown reasons | 0 | ? | ? | ? | |
| 8 | lower | 0/0/3<br>4<br>15 | 0 | 1 | 0 | 1 | 1 pupil in isolation: infected by a teacher not related to the regular school, whole class tested PCR- | 0 | 0 | 0 | ? | |
| | middle | 1/0/3<br>12<br>8 | 0 | 1 | 0 | 4 | 1 pupil in isolation: infected by a teacher not related to the regular school.<br>3 pupils in quarantine at unrelated time points: n=1 due to PCR+ HM, n=2 of for unknown reasons | 0 | 0 | 1 | 0 | |
| 9 | middle | 2/0/3<br>8<br>11 | 0 | 1 | 0 | 6 | 1 pupil in isolation: PCR+ during quarantine due to a PCR+ HM.<br>6 pupils in quarantine all due to PCR+ HM | 0 | 0 | 1 | 1 | Parallel class not participating in this study in quarantine |

| | | | | | | | | | | | | |
|---|---|---|---|---|---|---|---|---|---|---|---|---|
| | middle | 0/1/4<br>1<br>18 | 0 | 0 | 0 | 2 | 2 pupils in quarantine at unrelated time points due to PCR+ HM | 0 | 0 | 1 | 0 | due to infected teacher and class assistant |
| 10 | lower | 1/0/5<br>9<br>9 | 0 | 0 | 0 | 5 | 1 pupil in isolation due to PCR+ HM.<br>4 pupils in quarantine: n=3 each due to PCR+ HM, n=1 due to PCR+ contact | 0 | 0 | 1 | 1 | |
| | lower | 0/0/3<br>12<br>8 | 0 | 0 | 0 | 3 | 3 pupils in quarantine: n=1 due to PCR+ teacher not related to the regular school, n=2 both due to PCR+ HM of one of the 2 pupils, with whom both had close contact | 0 | 0 | 1 | 1 | |

HM – household member.

* T1+ denotes the number of seropositive pupils at T1 (June-July 2020); T2+ denotes the number of newly seropositive pupils at round T2 (October-November 2020); T3+ denotes the number of newly seropositive pupils at T3 (March-April 2021); *negative* denotes the number of seronegative pupils at round T3; *missing* denotes previously non-seropositive pupils who were not tested at T3. Total class size can be calculated by the sum of T1+, T2+, T3+, negative, and missing.

** Dates reported denote month and day(s). September to December refer to the year 2020, January to April – to the year 2021.

*** Possible index cases of individual diagnosed or suspected (RT-PCR+ or quarantined) children in the class.

**** Transmission among pupils/teachers in class was considered possible (1) when the time between RT-PCR diagnoses or quarantine start dates was 2 weeks or below, and not probable (0) if longer. Transmission was defined as unclear (?) if no or only a single child was diagnosed or quarantined in the class according to the interview information.
