## Appendix 5 for "Evolution of SARS-CoV-2 seroprevalence and clusters in school children from June 2020 to April 2021 reflect community transmission: prospective cohort study *Ciao Corona*"

**Appendix 5** Detailed results of survey for participants and non-participants on reasons for participation and non-participation

### Methods

The first two testing rounds of the prospective cohort study *Ciao Corona* took place in June-July 2020 (T1) and in October-November 2020 (T2). Testing rounds included blood sampling in participating children and adolescents aged 6-16 years and questionnaires completed by their parents. From conception of the study to the start of the recruitment of schools and children, we had a tight two months' window. The details of the study design are provided in the study protocol [1].

During the third round of testing between March 15 – April 16, 2021 (T3) parents of participants of the *Ciao Corona* study responded to an open question about reasons of participation, integrated in the regular follow-up questionnaire. Likewise, we assessed reasons for non-participation in the study by distributing a questionnaire to non-participating children and adolescents in the classes invited to participate in *Ciao Corona* study during T3. In the following sections we report findings from these two separate assessments to document and interpret reasons for participation (Section 1) and non-participation (Section 2) in our study.

### 1. Survey among participants of the Ciao Corona study – Reasons for participation

At the time of the third testing round in March 15 – April 16, 2021 (T3), we distributed a questionnaire that contained an open question about reasons for participation in the *Ciao Corona* study: “Why did you and your child decide to participate in the Ciao Corona study? Please give the most important reasons.” Two researchers (PA, SK) independently categorized the answers into categories related to “support of society and/or research”, “personal/family reasons”, or both. Agreement for discordant ratings was obtained by discussion.

**Results**

Overall, parents of 1356 from 2978 children participating in any of the testing rounds (46%) responded to the open question about reasons for participation in the study, of which 1331 (98%) were categorizable. As documented in Figure S1, the majority of survey respondents stated that they wanted to contribute to a better understanding of the severe acute respiratory syndrome coronavirus 2 (SARS-CoV-2) pandemic and help society to overcome the pandemic and/or support research (n=1192, 90%). 374 (28 %) expressed personal or family interests to learn if they had SARS-CoV-2 antibodies, but only in a minority (n=139, 10%) reported this as the only reason.

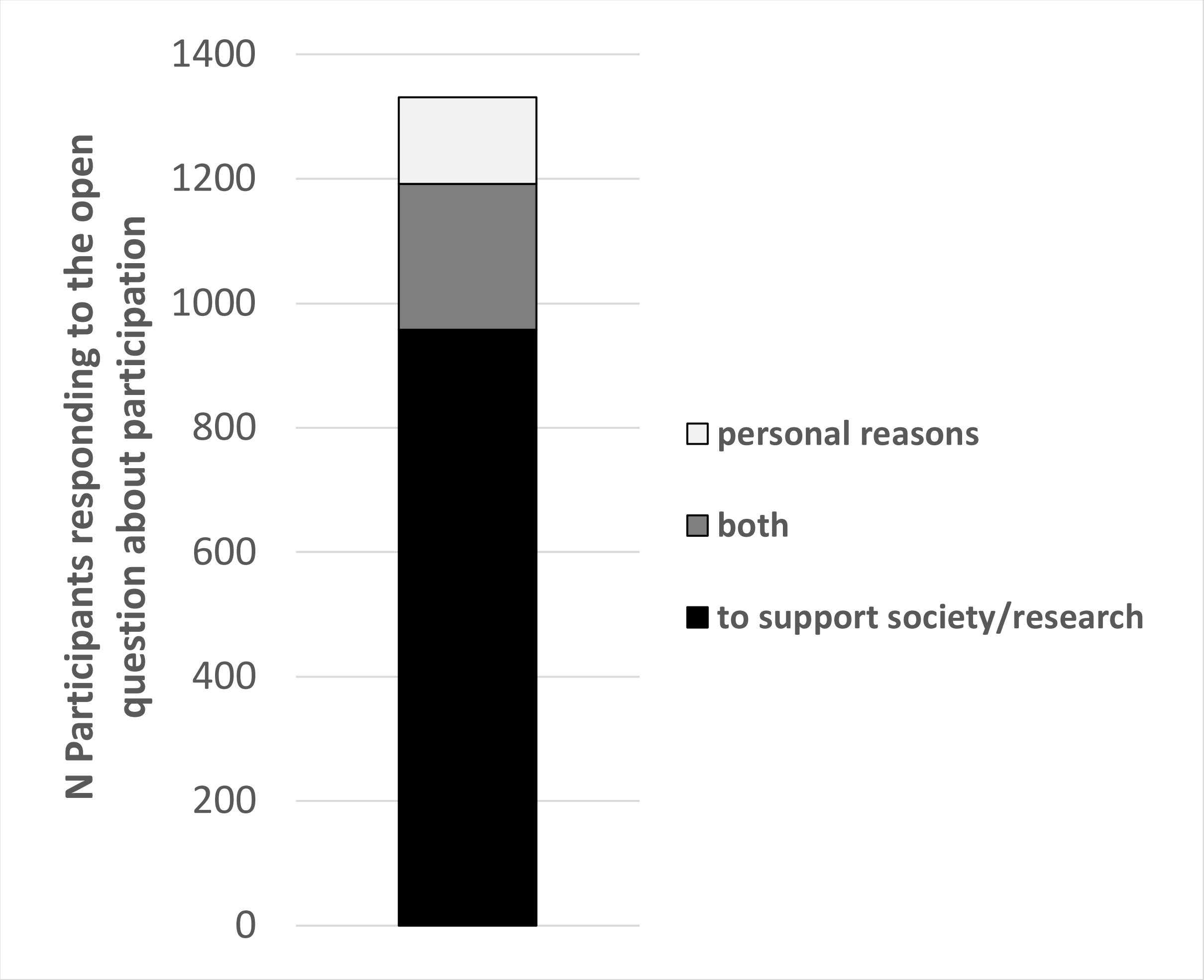


**Figure 1S** Reasons for participation in the Ciao Corona study among 1331 (45% of all) participants assessed at the time of the third round of testing in March/April 2021. Individuals who provided a categorizable comment to the open question why they took part in the study are included in this analysis.

Examples of individual comments are provided in the table below:

| **Society and/or research related reasons** |
|---|
| • "Without studies on Corona, there is no basis for fighting the virus"<br>• "We want to support research"<br>• "Because it is important that as many as possible participate so that the study is representative"<br>• "We think it is important to explore the role of children in the Corona pandemic"<br>• "That the virus will go away quickly"<br>• "The more data and studies we have, the better we will come out of this situation and the better policy decisions will be supported" |

| |
|---|
| • “I think the study makes sense. My son asked me. I am convinced that we make better progress with facts than with assumptions”<br>• “That we can contribute to scientific research on the virus, which hopefully will help to find a solution in the near future”<br>• “Important for the general public”<br>• “We wanted to help“<br>• “To use all channels together to fight the pandemic. To know if we have gone through the disease unnoticed“ |
| **Personal/family reasons** |
| • “Personal curiosity about the development of the situation. The promised goodies were also quite motivating for my child”<br>• “My daughter wanted to take part”<br>• “To protect the grandparents, and because this study is important”<br>• “That we can get tested for free”<br>• “Because there is a gift!” |

**Interpretation of individual reasons for participation in *Ciao Corona***

90% of families reported an “altruistic reason” of participation in the Ciao Corona study, for example, the wish to support research to learn more about the novel coronavirus (SARS-CoV-2) or to help society to overcome the pandemic. Participation because of personal reasons and benefit, such as getting the individual SARS-CoV-2 antibody result, was not a dominant reason.

Data on participation rates for school-based interventions and prevention programmes are generally scarce and often not reported in the original studies.[2] A literature review including 481 school-based studies revealed that only 11.5% of studies reported both consent procedures and participation rates. In studies using active consent procedures by parents, like in the Ciao Corona study, the mean participation rate was 65.5% (range: 11–100%), but most of these studies focused on questionnaires or lifestyle changes and did not include blood sampling.[2]

We achieved a comparable participation rate (of about 50%) of children and adolescents to that of school-based intervention study including blood sampling, reported by Group et al.[3] However, this large study had a much higher attrition rate (defined as the proportion of enrolled participants not attending the last follow-up) than our study (30% versus 16%). This randomised controlled trial in school-aged children included only grade 6 children (on average older than in our study) predominantly from deprived populations. Study investigators paid 50-60 $ for children and 35$ for parents as an incentive for their participation in each of the two health assessments rounds over 2 years. Recruitment and retention rates reported in other school-based studies were much lower than in our study, despite the fact that financial incentives to schools and parents and children were provided.[4]

School-based studies benefit from being conducted in a location familiar to the child and with precious peer support, which may mitigate mistrust in research and reduce barriers of research in hospital settings.[5-7] On the other hand, they are complex in many other respects (e.g., convincing school principals, language barriers, additional time investment for school staff, and potential conflicts with compulsory school lessons). Many hurdles need to be overcome by a whole chain of agreements with cantonal authorities, ethical committees, school authorities, individual school principals and teachers, children and families – until the written parental consent is obtained. Despite the urgency of setting-up our study within two months, the team invested substantially in communicating with school principals, preparing multi-facetted study materials including a website (www.ciao-corona.ch) with child-friendly pictures and videoclips explaining the testing procedures to children and their parents. Further, we organised online meetings for school personnel and parents to explain the rationale of the study, goals, testing procedures, and offered time to discuss individual questions. We also gave children, parents and school staff the possibility to get into contact with us any time by telephone hotline and email.

It is important to highlight that retention of study participants remained very high (89 % at T2, 84% at T3) over the course of the study, including three testing phases over 10 months. The course of the SARS-CoV-2 pandemic itself with high infection rates at the community level, the report of individual and school test results to study participants after each testing phase, involvement of various stakeholders (e.g., school principals, department of education of the canton of Zurich), and the investment in different communication channels (e.g., study website, social media). Also, the  frequent media presence, often in collaboration with the cantonal school authorities, may have contributed to the high participation rate in the Ciao Corona study.

### 2. Survey among non-participants of the Ciao Corona study – Reasons for non-participation

During the third testing round at schools (T3), we distributed an anonymous paper questionnaire to children of the invited classes that did not participate in the T3 Ciao Corona assessment to elucidate reasons for non-participation. Parents of non-participating children were given the opportunity to either complete the questionnaire online (via a public survey link using REDCap, Research Electronic Data Capture, Vanderbilt University, US), or on paper (to be sent back with a pre-paid mailing envelope).

The questionnaire was available in 10 languages (German, English, French, Italian, Spanish, Portuguese, Tamil, Turkey, Croatian, and Albanian) and contained six questions: 1) sex (male, female, prefer not to say), 2) name of the school, 3) school level (lower, middle, upper school), 4) whether the parents were contacted in the past to participate in the Ciao Corona study (yes, no), 5) a list of predefined reasons for non-participation followed by a free text field for comments (Of note, multiple answers could be given for 12 pre-defined reasons and one additional field for "other reasons") , and 6) who in the family made the decision not to participate (parents, child, both together).

We then categorized individual comments by their general emotional connotation towards the study into a more "negative", "positive", or "neutral" response, and whether non-participation reasons were potentially modifiable by the study team.  Two members of the study team screened and categorized individual responses independently (SK, TR). Discrepancies categorizations were discussed and consensus achieved though discussion.

#### Results

50/55 schools agreed to the distribution of the questionnaire within participating classes. At the testing day, 5001/5205 (96%) children in the classes invited to the study in these schools were present, and 2342 of them (47%) participated in T3 testing round. From the remaining 2659 children eligible for the non-participation survey, parents of 712 (27 %) children responded. Of those, 328/695 (47%), 192/695 (28%), and 175/695 (25%) of parents responded for children from lower, middle, and upper school level (17 did not report school level). Among survey participants, 673 (94.5%) responded in German, 12 (1.7%) in English, 7 (1%) in Albanian, 6 (0.8%) in Italian, 5 (0.7%) in Turkish, 4 (0.6) in Portuguese, and 2 (0.3%) in

Spanish. 591/657 (90%) participants reported that they received an invitation to take part in the study and 66 (10%) denied that they received an invitation (55 did not respond to this question). Those who reported an individual comment that their child participated in T1 or T2 were removed from the analysis (n=28, 4%).

The final decision not to take part in the study was made by the parent(s) alone for 64/544 children (12%), parent(s) and child together for 205 (38%), or the child alone for 275 children (51%) (20 parents did not respond to this question). **Figure 2S** (also presented in the main manuscript) provides an overview about reasons for non-participation according to school level. The most frequently reported reason for non-participation was fear of blood sampling (358/564, 64%), which was reported more frequently among girls (189/276, 69%) compared to boys (158/259, 61%), and in children from lower school level (191/264, 72%) compared to middle (85/153, 56%) and upper school level (79/141, 56%), respectively.

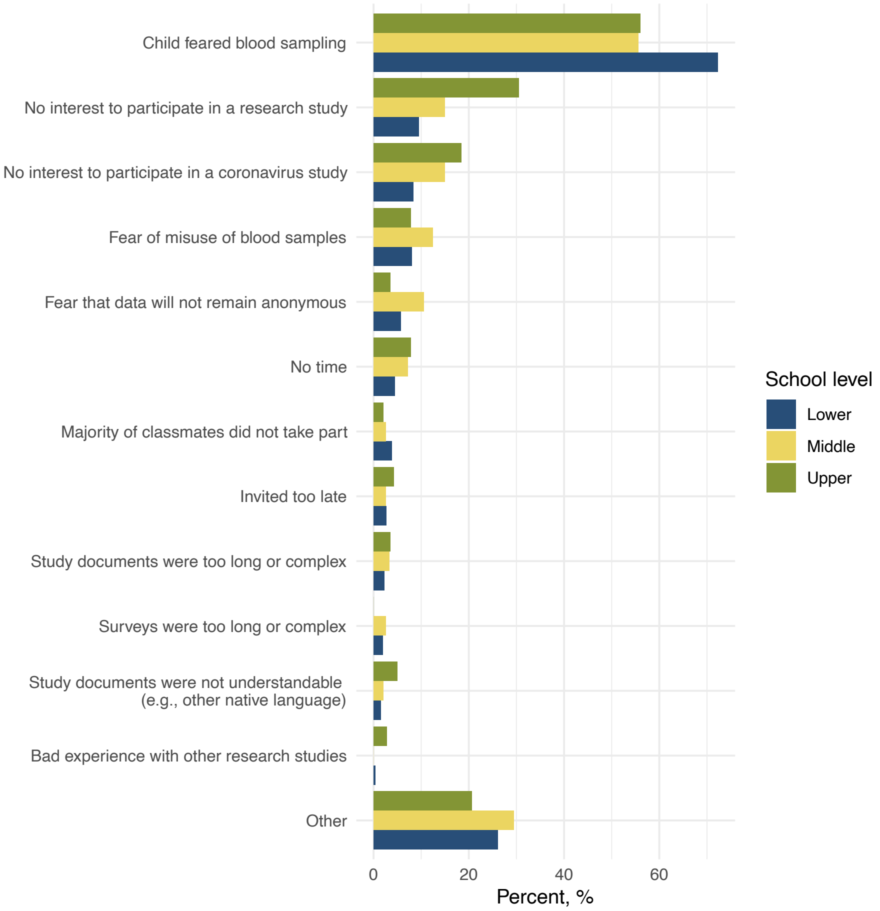


**Figure 2S** Reasons for non-participation in the Ciao Corona study assessed during the third round of assessment in March/April 2021 among 567 eligible participants according to the school level (e.g. lower=grades 2-3, middle=grades 5-6, upper=grades 8-9). Multiple answers were allowed for the 13 pre-defined reasons including a field for “other reasons”

341/684 survey participants (50%) provided individual comments that we categorized into "overall negative", "overall positive", or "neutral" responses, and modifiable versus non-modifiable (**Table 1S**)

**Table 1S** Emotional connotation and modifiability of reasons for non-participation among 341/684 survey respondents providing specific comments

| | **N (%)** |
|---|---|
| *Emotional connotation of comment* | **N = 341** |
| Overall negative | 101 (30) |
| Overall positive | 92 (27) |
| Neutral | 146 (43) |
| Unclear | 2 (0) |
| *Modifiability of reasons* | |
| Non-modifiable | 266 (78) |
| Modifiable | 53 (16) |
| Unclear | 22 (6) |

Individual comments from survey participants to the question "What could we do differently so that you would participate in the Ciao Corona study?" are given in the table below:

| **Overall negative – non-modifiable** |
|---|
| • "Why don't you do such studies on adults rather than on children/adolescents? Especially if no one in the class has had Corona, an antibody study is probably not that useful".<br>• "I am absolutely against the whole testing thing. If we stopped, the whole Corona craze would be over. Tests have been proven to be unreliable (see WHO)".<br>• "Because we feel that the current situation and the way in which research is being carried out is not honest. The Federal Council is not interested in scientific facts anyway!"<br>• "Because we didn't understand the point of starting a study with children. We felt it was pointless. We do not support this "scaremongering" and want to be left alone with studies and everything else that has to do with Corona".<br>• "Not at all – Corona lie" |

| |
|---|
| • "Don't write in such a complicated way" |
| **Overall negative – potentially modifiable** |
| • "It was unclear what happens to the data and what is done with the blood. Pass on/sell to third parties?!"<br>• "With better communication. Clear formulation of hypotheses. Unfortunately, it gave the impression in the first test that people simply want to make a name for themselves with the topic and that the children at the schools are the easiest to access in order to have test results. That bothered me a lot".<br>• "A positive test result would have even more consequences for our self-employment. Fear of existence, shop closure, loss of work".<br>• "No! Many studies are funded by pharmaceutical companies, and I don't want to participate to support the pharmaceutical lobby". |
| **Overall positive – non-modifiable** |
| • "With a miracle cure for fainting during blood sampling"<br>• "Our son collapses when blood is taken, cannot be mobilised (stand up) for up to four hours afterwards and needs medical supervision each time. Therefore, he could unfortunately not participate" |
| **Overall positive – potentially modifiable** |
| • "Better information about blood collection"<br>• "No needles - difficult to implement. The anaesthetic patch should have been promoted more"<br>• "Participation of parent during blood collection"<br>• Eventually my child would have taken part if we parents could have been present. But this was not possible due to Corona" |
| **Neutral – non-modifiable** |
| • "Fear of being quarantined as an extended family"<br>• "Taking blood from the finger, finger prick" |
| **Neutral – potentially modifiable** |
| • "Anonymized data (not just encrypted, with the explicit option to de-encrypt on demand)"<br>• "The teacher could have explained the study to the non-speaking German parents" |

#### Interpretation of individual reasons for non-participation in *Ciao Corona*

Overall, 27% of eligible respondents participated in this anonymous survey and provided reasons for non-participation in the Ciao Corona study. This information is helpful to better understand the huge problem of non-participation and selection bias in virtually every population-based study. The most frequent reason for non-participation was fear of blood sampling (64%), which is understandable given the fact that we invited as young as 6 years old children. Even with a videoclip available explaining the procedures and the use of a plaster with an anesthetic cream to numb the skin over the blood sampling place, this fear was still prominent. Yet, we do not know how many of these "anxious" children did indeed watch the videoclip although a lot of teachers reported that they did so even in class. We could possibly have put even more effort into explaining the blood sampling procedures, for example by preparing more precise video-clips with each step included and explained (meeting in the classroom, application of patches with an anaesthetic cream on the location of venipucture, blood drawing procedures, selection of a plaster, get a sweet and a present) that could be watched in the classroom or at home. We could have found a way to always allowing a parent to be present for the blood sampling (which was not always the case due to school-based mitigation strategies), especially in the younger age group, e.g., by testing the child with a present parent in a bus outside the school.  At least, we could have demonstrated the blood sampling on the classroom teachers. From a logistic perspective, most of these adaptations would have been a major challenge, if not even an unrealistic hurdle to take.

Other reasons included lack of interest in research studies in general or SARS-CoV-2-related research (16% and 13%). About 16% reported concerns about ethical issues, e.g., "experimenting" with the child's blood or lack of anonymity. Although a minority, parents with such concerns have to be taken seriously. Although we aimed to provide clear statements in the information and consent sheets that all data got fully anonymised and the blood was used for corona-related research, more effort might have been needed.

Finally, about half of the survey participants provided individual comments and reasons for non-participation in the study. Of those, 70% were either positive or neutral, whereas 30% were categorized as negative. The vast majority of reported individual reasons for non-participation were non-modifiable due to study design (e.g., need for venous blood sampling). About 16% of responses were categorized as modifiable. Most of those comments were very useful for future study planning: they addressed the communication with parents to rule out misinformation/misinterpretation of study contents, the wish for more and repeated information, more simplified explanations of the study objectives and planned testing procedures. Yet, the latter is often interfering with the requirements of the ethical committees requesting an extensive and detailed study information regarding the aim of the study, eligibility criteria, general information about the study, detailed procedures, benefit for participants, rights and obligations, measures taken, confidentiality of data and probes, compensation, insurance coverage, funding, etc. Future collaborative work with ethical committees, schools, children and parents themselves are needed to improve communication strategies that could further increase participation.

Despite a high participation rate of almost 30%, selection bias among eligible survey participants is still expected and may hamper generalisability of the survey. We did not collect socioeconomic characteristics with this anonymous survey, but the fact that fewer than expected parents responded in other languages than German suggest selection into the study. This could signal differential study participation with less disadvantaged families participating more frequently, as was observed in other studies focusing on children[8 9] which could limit the generalisability of our study findings. Although likely not possible to remove complete, this challenge could be addressed by optimising communication by establishing emotional bonds to the eligible population, simplifying and visualising study information, providing repeated information and communicating the value of each individual participant beyond simply providing material and financial incentives.
